# The periaqueductal grey in chronic low back pain: dysregulated metabolites and function

**DOI:** 10.1101/2023.06.01.23290820

**Authors:** Laura Sirucek, Iara De Schoenmacker, Lindsay Gorrell, Robin Lütolf, Anke Langenfeld, Mirjam Baechler, Brigitte Wirth, Michèle Hubli, Niklaus Zölch, Petra Schweinhardt

## Abstract

Mechanisms underlying chronic pain are insufficiently understood. Preclinical evidence suggests a potential contribution of excitatory glutamatergic and inhibitory GABAergic imbalances in pain-relevant brain areas, such as a lower excitatory/inhibitory tone in the brainstem periaqueductal grey (PAG). This cross-sectional magnetic resonance spectroscopy (MRS) study investigated whether a lower excitatory/inhibitory tone is also observed in the PAG of patients with non-specific chronic low back pain (CLBP) and whether this would relate to altered psychophysical measures of descending pain modulation and experimental pressure pain sensitivity. Specifically, the ratio between pooled glutamate and glutamine and GABA levels (Glx/GABA), Glx and GABA in the PAG were compared between CLBP patients and pain-free controls. Further, associations of Glx/GABA with conditioned pain modulation (CPM) effects and pressure pain thresholds (PPTs) were assessed.

MRS was acquired on a 3T Philipps MR system using a point-resolved spectroscopy sequence optimized with very selective saturation pulses (OVERPRESS) and voxel-based flip angle calibration in a 1.1 mL volume of interest. Data from 41 CLBP patients (median [interquartile range]: 54 years [41 - 65], 22 females) and 29 age- and sex-matched controls (47 years [34 - 67], 17 females) fulfilled MRS quality criteria. CPM and PPTs were assessed at the lower back as most painful area and the non-dominant hand as pain-free control area. The CPM paradigm consisted of PPTs applied before, during (parallel CPM effect) and after a cold water bath and an ambient temperature water bath as control paradigm to identify ‘true’ CPM effects.

In the PAG of CLBP patients, a lower Glx/GABA ratio, i.e. a lower excitatory/inhibitory tone, was observed (*P* = 0.002, *partial η^2^* = 0.14) driven by decreased Glx (*P* = 0.012, *partial η^2^* = 0.11) and increased GABA (*P* = 0.038, *d* = 0.46). CLBP patients showed disrupted associations between Glx/GABA and PPTs compared to controls in both areas (lower back: *P* = 0.004, *partial η^2^* = 0.12; hand: *P* = 0.002, *partial η^2^* = 0.16). In controls, lower Glx/GABA was associated with lower PPTs (lower back: *r* = 0.48, *P* = 0.009, hand: *r* = 0.53, *P* = 0.003), but this link was missing in CLBP patients (*r’s* > -0.23, *P’s* > 0.150). Additionally, CLBP patients with more severe clinical pain showed smaller CPM effects at the hand (*rho* = 0.54, *P* = 0.003).

These findings suggest a dysfunction of the PAG in patients with CLBP and might indicate altered descending inhibition of deep tissue afferents.

## Introduction

Acute pain is vital to protect organisms from harm.^1^ However, when pain becomes chronic, it often loses its protective function and causes major suffering for the individual^2^ and tremendous societal costs.^3^ Effective treatment options are lacking because the mechanisms underlying the development or maintenance of chronic pain are insufficiently understood. Preclinical evidence suggests excitatory glutamatergic/inhibitory GABAergic imbalances in pain-relevant brain regions as a potential contributor to chronic pain.^4^ In chronic pain patients, alterations of glutamate,^5–13^ glutamine,^14–16^ pooled levels of glutamate and glutamine (Glx),^5–7,9–13,17–26^ or GABA^27–35^ have been demonstrated in pain-relevant brain regions, including the insula,^10, 14–17, 19, 30^ cingulate cortex,^5, 6, 8, 9, 11, 12, 16, 21, 27, 32^ or thalamus^18, 26, 31, 33^ using proton magnetic resonance spectroscopy (^1^H-MRS).^36^

One brain region in which excitatory/inhibitory imbalances might have a substantial impact on pain processing is the periaqueductal grey (PAG). This key descending pain modulatory brainstem region^37^ exerts pain inhibition or facilitation via descending projections to the rostral ventromedial medulla (RVM) and the spinal cord.^38^ Based on the ‘GABA disinhibition’ hypothesis,^37^ PAG-driven descending pain inhibition is activated by switching off tonic GABAergic controls of descending glutamatergic projections. Conceivably, an excessive GABAergic tone in the PAG might impede its inhibitory function and contribute to aberrant pain processing. A reduced glutamatergic tone could have a similar effect because glutamate release can indirectly inhibit GABAergic controls in the PAG.^39^ In line with these notions, an augmented GABAergic tone^40^ as well as a hypo-glutamatergic state^40^ has been observed in the PAG of animal models of chronic neuropathic pain.^41^ In humans, investigations of PAG neurochemistry are scarce,^42–45^ possibly because ^1^H-MRS in the PAG is technically challenging. The PAG’s small size of approximately 4-5 mm (diameter) x 14 mm^46^ makes it difficult to obtain sufficient signal. Additionally, its proximity to pulsating vessels and CSF causes high levels of physiological noise,^47^ further decreasing the signal-to-noise ratio (SNR).^48^ Existing PAG ^1^H-MRS studies have addressed these challenges by using long echo times which reduces the influence from macromolecules and lipids on the spectrum benefitting reliable metabolite detection but hampers detection of J-coupled metabolites such as Glx or GABA,^43^ or by using large volumes of interest (VOI), which increases SNR but limits regional specificity.^42, 45^

In the present study, we adopted an improved approach enabling high-quality ^1^H-MRS acquisition in the PAG (https://doi.org/10.1101/2023.03.29.534678) with the aim to investigate neurochemical alterations in patients with non-specific chronic low back pain (CLBP). In brief, Glx and GABA were measured in a 1.1 mL small VOI using a Point-RESolved Spectroscopy sequence (PRESS)^49, 50^ combined with Very Selective Saturation (VSS) pulses (OVERPRESS)^51–53^ and voxel-based flip angle calibration^54, 55^ to optimize MRS acquisition, and with spectral registration to optimize MRS preprocessing.^56^ Specifically, the PAG’s excitatory/inhibitory balance, conceptualized as Glx/GABA, was compared between patients with CLBP and pain-free controls. Lower Glx/GABA, i.e. decreased Glx and/or increased GABA, was expected in CLBP patients. In addition, it was investigated whether Glx/GABA was related to a psychophysical measure of descending pain modulation, i.e. conditioned pain modulation (CPM).^57^ CPM is considered the human counterpart of diffuse noxious inhibitory controls (DNIC)^58, 59^ and assesses how the perceived pain intensity of a noxious test stimulus is altered by another, heterotopically applied, noxious “conditioning” stimulus (CS). A lower excitatory/inhibitory tone, i.e. lower Glx/GABA, was expected to be associated with smaller CPM effects indicative of weaker descending pain modulatory capacity. Finally, associations of Glx/GABA with experimental pressure pain sensitivity, and associations of Glx/GABA and CPM effects with clinical characteristics were explored.

## Materials and methods

### Participants

CLBP patients were consecutively recruited via the Balgrist University Hospital and advertisements in Swiss Chiropractic practices and patient magazines. Individually age- and sex-matched, pain-free controls were recruited via online advertisements and oral communication. The recruitment period, data collection period and sample size calculation are described in Supplementary Methods M1. Inclusion criteria for CLBP patients were between 18 and 80 years of age, good general health and CLBP as primary pain complaint without “red flags” (e.g. signs of infection, fractures, inflammation) and of a duration longer than 3 months. For controls, the same inclusion criteria were applied with the additional requirement that they had not experienced low back pain lasting longer than 3 consecutive days during the last year.

Exclusion criteria for both cohorts comprised any major medical or psychiatric condition other than CLBP, symptomatic radiculopathy (i.e. motor and sensory deficits), pregnancy, inability to follow study instructions or contraindications to MRI. The study was approved by the local ethics committee “Kantonale Ethikkommission Zürich” (Nr.: 2019-00136), registered on clinicaltrials.gov (NCT04433299), and performed according to the guidelines of the Declaration of Helsinki (2013). Written informed consent was obtained from all participants before the start of the experiment.

### Study design

This study was part of a larger project (Clinical Research Priority Program “Pain”, https://www.crpp-pain.uzh.ch/en.html) which comprised 3 experimental sessions of approximately 3 hours each and electronic questionnaires. In the first 2 sessions, participants underwent clinical, neurophysiological and psychophysical assessments. The CPM assessment was always performed in the second session preceded by the acquisition of pain-evoked potentials. During the third session, participants underwent 2 MR measurements, 1 ^1^H-MRS scan and 1 resting state functional MRI scan with a break of 1 hour between. All scans were performed after 12 pm. The present study concerns the ^1^H-MRS, the CPM, the pain drawings and experimental pressure pain sensitivity of the first session’s psychophysical assessment and questionnaire data. Fig. 1 shows an overview of the study design.

**Figure 1.**
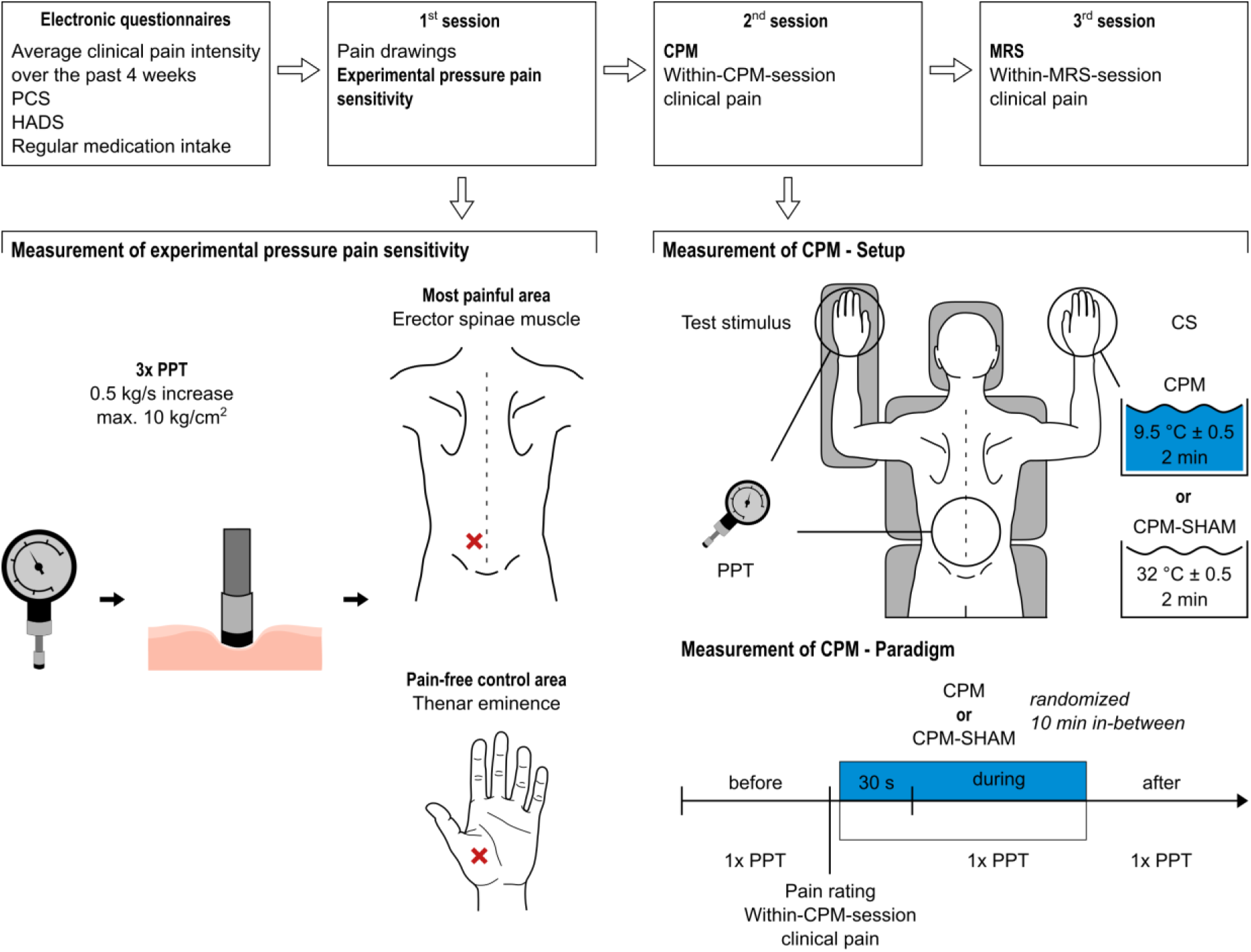
Study design overview. Main outcomes of interest are highlighted in bold italic. Red crosses indicate locations at which PPTs were assessed, i.e. over the erector spinae muscle within the lower back (most painful area) and at the thenar eminence of the non-dominant hand (pain-free control area). The CPM-SHAM paradigm was only performed at the non-dominant hand.

### Magnetic resonance spectroscopy

Substantial material in this section is identical to a technical note (https://doi.org/10.1101/2023.03.29.534678) focused on methodological aspects of high-quality ^1^H-MRS acquisition in the PAG using the data of the controls. Technical details of the ^1^H-MRS are listed in Supplementary Table 1. The following sections provide a brief overview of the applied methods.

### Acquisition

^1^H-MRS was acquired on a 3T MR system using a 32-channel head coil (Philips Healthcare, Best, The Netherlands). Prior to ^1^H-MRS acquisition, high-resolution (1 mm^3^ isotropic) anatomical T_1_-weighted images were obtained (acquisition time: 7 min 32 s). Based on the 3D T_1_ images, the VOI was placed to cover the PAG according to anatomical landmarks by the same examiner (LS) for all participants (Fig. 2A). Spectra were localized using a water-suppressed single-voxel OVERPRESS sequence^49–53^ (repetition time (TR): 2500 ms, TE: 33 ms, number of signals averaged: 512 divided into 8 blocks of 64; acquisition time: 23 min 20 s). The use of VSS pulses minimizes errors in chemical-shift displacement and allows for consistent localization volumes across all metabolites of interest (Fig. 2A). The voxel-based flip angle calibration^54, 55^ achieves an optimal flip angle within the VOI. Accounting for the VSS pulses, the resulting VOI size was 8.8×10.2×12.2 mm^3^ = 1.1 mL. For each individual, 2 water signals were acquired: 1 obtained from interleaved water unsuppressed spectra (1 before each of the 8 blocks) for eddy-current correction and internal water referencing corrected for partial volume and tissue-specific relaxation effects using literature-based T_1_ and T_2_ relaxation times;^60^ 1 measured in a separate water reference scan after the ^1^H-MRS acquisition in the PAG within the same VOI with a TR of 10000 ms and varying TEs (33/66/107/165/261/600 ms), allowing estimation of the T_2_ relaxation time of water within the VOI. Hereby, a literature-independent, subject-specific approximation of the fully-relaxed water signal was obtained.^61^

**Figure 2.**
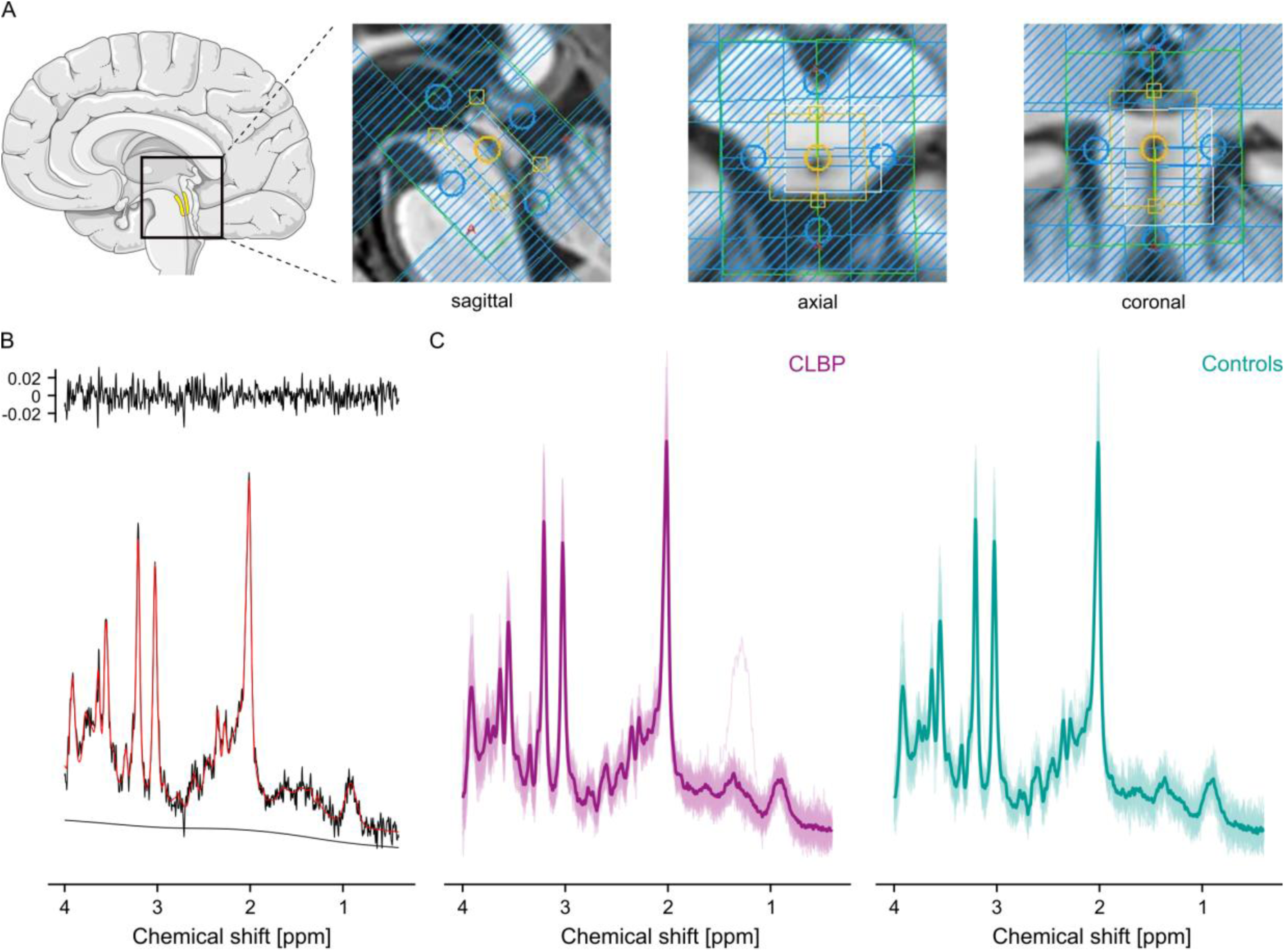
^1^H-MRS VOI placement and acquired spectra. (**A**) The VOI was placed according to anatomical landmarks such as the cerebral aqueduct. The yellow box represents the nominal size of the VOI (11×15×18 mm^3^ [APxLRxFH]). Blue-shaded areas represent the VSS bands used to minimize errors in chemical-shift displacement and to achieve consistent localization volumes across all metabolites of interest. The unshaded area within the yellow box represents the final VOI size (8.8×10.2×12.2 mm^3^). (**B**) Representative single spectrum (SNR = 19, FWHM H_2_O = 5.1 Hz, age = within 53.5 years (median age of the cohort) ± 5). (**C**) Overlaid single spectra together with the group average (bold) for CLBP patients (purple) and controls (turquoise). The schematic brain was adapted from Servier Medical Art (smart.servier.com).

### Preprocessing and analysis

The T_1_-weighted images were segmented into grey matter (GM), white matter (WM) and CSF to obtain the respective tissue fractions (in %) within the VOI using SPM12.^62^

Frequency alignment was performed using spectral registration in the time domain (adopted from^56, 63^). For that, the data were filtered with a 2 Hz Gaussian filter. Only the first 500 ms were used for alignment and the single averages were aligned to the median of all averages. Preprocessing was performed by an author blinded to the hypotheses of the study (NZ).

The preprocessed spectra were inspected for artefacts. Spectra with artefacts were excluded from further analyses, as were spectra presenting with insufficient quality,^55^ i.e. with a full width at half maximum (FWHM) value of the unsuppressed water peak (FWHM H_2_O; shim quality indicator) above 2.5 median absolute deviation (MAD)^64^ of the group median or with a SNR below 2.5 MAD of the group median.

### Metabolite quantification

Spectra were analyzed using LCModel (6.3)^65^ with a metabolite basis set simulated using FID-A taking into account spatial localization and actual radiofrequency pulse shapes.^63^ Metabolite concentrations are reported as ratios to the unsuppressed water signal and reflect an estimation of metabolite concentration in moles per kg of tissue water excluding water within CSF. As primary outcome, the subject-specific water signal was used. In addition to being independent of literature-based tissue-specific T_1_ and T_2_ relaxation times, known to vary across brain regions^66–68^ and possibly not generalizable to the PAG, this approach relies less on the correct segmentation of GM, WM, and CSF compared to the literature-based approach. For comparison, all analyses were also performed using the literature-based water signal.

### Conditioned pain modulation

Participants were tested in a prone position in a quiet room (temperature 20-25 °C). The CPM assessment was performed by a trained experimenter blinded to the hypotheses of the study. Participants were given standardized verbal instructions.

CPM was assessed at the patient’s most painful area in the lower back and at the non-dominant hand as a remote, pain-free control area. The order of the 2 areas was randomized and counterbalanced. The exact area on the lower back, the laterality of the hand (right or left) and the order of the 2 areas in each control was identical to the patient to whom they were individually age- and sex-matched.

The CS consisted of a circulating cold water bath (9 °C ± 0.5) in which the participants immersed their dominant hand up to the wrist for 2 min. The perceived pain intensity of the CS was rated on a numeric rating scale (NRS) from 0 ‘No pain’ to 10 ‘Maximum pain tolerable’ directly after immersion, 30 s after immersion and immediately before withdrawal of the hand. If the pain became intolerable, participants were allowed to withdraw their hand but were encouraged to re-immerse their hand as soon as possible.

The test stimulus consisted of pressure pain thresholds (PPT) assessed before, during (30 s after CS onset; ‘parallel CPM effect’), and after (mean 2 min 58 s, SD = 49 s after CS end; ‘sequential CPM effect’) the CS. PPTs were chosen as test stimulus because (1) deep afferents are likely to play a more important role compared to superficial afferents in CLBP as musculoskeletal pain condition and (2) the combination with a cold water bath as CS has been shown to be the CPM paradigm with the highest intra-session reliability^69^ and with larger CPM effects compared to other TS-CS combinations.^70^ Pressure was applied using a hand-held mechanical algometer (Wagner Instruments, Greenwich, CT, USA) with a circular rubber tip (1 cm diameter). For the lower back, PPTs were assessed over the erector spinae muscle at the level and the body side of the most painful area. For the hand, PPTs were assessed at the thenar eminence. One PPT per timepoint was determined using the method of limits.^71^ If no pain was reported at 10 kg/cm^2^ (safety cut-off to avoid tissue damage), a value of 11 kg/cm^2^ was assigned as the PPT. Three additional test stimuli were administered, i.e. pressure temporal summation of pain (TSP), heat pain thresholds and heat TSP. The order of pressure and heat assessments was randomized and counterbalanced and TSP was always performed after the threshold assessments. These test stimuli answer a different research question and will not be discussed further; relevant here is that the paradigm was identical between the CLBP patients and the controls.

At the hand, an additional CPM-SHAM paradigm was performed to test whether the used CPM paradigm induced a ‘true’ CPM effect beyond repeated-measures effects.^72^ The CPM-SHAM paradigm was performed identically to the CPM paradigm except for an ambient temperature water bath (32 °C ± 0.5) as CS. Within the assessment of the hand, the order of CPM and CPM-SHAM paradigms was randomized and counterbalanced with a minimum of 10 min between.

### Clinical characteristics

As part of the electronic questionnaires, CLBP patients reported their average clinical pain intensity over the past 4 weeks (NRS: 0 ‘No pain’ to 10 ‘Maximum pain’). To allow for differentiation between this ‘trait’ pain and the ‘state’ pain during the experimental sessions, 2 additional pain intensity ratings were acquired: (1) *within-MRS-session clinical pain*, where after the ^1^H-MRS scan, CLBP patients rated the maximum pain they had experienced during the scan (NRS: 0 ‘No pain’ to 10 ‘Most intense pain tolerable’) and (2) *within-CPM-session clinical pain,* where CLBP patients reported the current pain intensity in their most painful area before the first water bath on the same NRS as for (1).

CLBP patients completed pain drawings before the first experimental session to assess the spatial pain extent (% of total body area) of their typically painful body areas^73–76^ (Supplementary Methods M2).

Participants also completed the Pain Catastrophizing Scale (PCS)^77^ and the Hospital Anxiety and Depression Scale (HADS).^78^ The PCS consists of 13 items with a score range between 0 and 52 (> 30 clinically relevant level of catastrophizing). The HADS consists of 14 items with a 7-item anxiety-subscale and a 7-item depression-subscale, with a score range between 0 and 21 per subscale (8-10 moderate and 11-21 high probability for a mood disorder).

### Confounding factors

Prior to the CPM and the ^1^H-MRS session, information about the menstrual cycle phase was obtained if applicable, i.e. in premenopausal women not using menstruation-suppressing contraceptives.

In the electronic questionnaires, participants were asked to indicate any regular medication intake. Medications were classified according to the ATC/DDD classification by the World Health Organization (http://www.whocc.no/atc_ddd_index/). The following categories were considered pain-relevant: M01A (anti-inflammatory and anti-rheumatic drugs and non-steroids), N02 (analgesics), N03 (antiepileptics), N05 (psycholeptics), and N06 (psychoanaleptics).

### Statistical analysis

All statistical analyses were performed in RStudio^79, 80^ for Mac (2022.12.0+353). Statistical significance was set at *α* = 0.05 with a false discovery rate (FDR) correction per tested research question. The number of corrected tests per research question is indicated as *n*-FDR.

Depending on the statistical test used, raw values or model residuals were assessed for normal distribution using inspection of histograms and QQplots. All ordinal or non-normally distributed variables are reported as median (interquartile range) and were analyzed using non-parametric tests. All continuous variables are reported as mean (SD) and were analyzed using parametric tests.

Group comparisons were performed using linear models (Supplementary Methods M3). This allowed for the examination of the potential influences of age and sex on the dependent variables and of influential cases in a standardized manner (Supplementary Methods M4). For all models, the number of identified influential cases is indicated as *n*-IC. If removal of influential cases changed the statistical inferences, the results of the model without the influential cases are reported (indicated as *n*-IC^†^). Otherwise, the results of the full data set are reported.

Effect sizes are reported using *partial η^2^* (small: 0.01, medium: 0.06, large: 0.14)^81^ for linear models, Cohen’s d (small: < 0.5, medium: 0.5-0.8, large: > 0.8)^81^ for t-tests and r (small: 0.1-< 0.3, medium: 0.3-< 0.5, large: ≥ 0.5)^82^ for Wilcoxon rank-sum tests. No effect sizes are reported for linear mixed models because no agreement on standard effect sizes exists.^83^

### Magnetic resonance spectroscopy

Group comparisons were performed for: (1) Glx/GABA as main outcome of interest; (2) Glx and GABA separately to examine which metabolite drove the observed group difference in Glx/GABA (*n*-FDR: 2); and (3) tCre (creatine + phosphocreatine), tCho (glycerophospho-choline + phosphocholine), tmI (myo-inositol + glycine), tNAA (N-acetylaspartate + N-acetylaspartylglutamate), as well as GM, WM and CSF tissue fractions (only the analysis of Glx and GABA were included in the multiple comparison correction because the other outcomes were not part of the primary research question). For GABA and CSF tissue fraction, group comparisons were performed using a Welch’s t-test and Wilcoxon test, respectively, because linear model assumptions were not met.

Because differences in Glx or GABA might be confounded by differences in GM or WM tissue fractions, it was investigated whether Glx or GABA correlated with GM or WM tissue fractions in the controls, i.e. a ‘healthy’ state, using Pearson correlations (without multiple comparison correction to minimize the risk for false negatives).

### Conditioned pain modulation

CPM effects were calculated as follows: parallel CPM effect = PPT before – PPT during, sequential CPM effect = PPT before – PPT after. Thus, inhibitory CPM effects are denoted by a negative value and facilitatory CPM effects by a positive value.^84^ For all *within-subject* analyses, absolute values were used. For all *between-subject* and correlational analyses, relative differences, i.e. ((PPT before – PPT during) / PPT before) * 100, were used to allow direct comparisons between the lower back and the hand (which might have different PPTs). Maximum CPM inhibition and CPM facilitation was set to ±100%.

#### Within-subject analyses

Firstly, the presence of ‘true’ CPM effects beyond repeated-measures effects was tested using a linear mixed model on the data of the controls (Supplementary Methods M3). The effect of interest was the interaction between ‘timepoint’ (levels: ‘before’, ‘during’, and ‘after’) and ‘paradigm’ (levels: ‘CPM’, ‘CPM-SHAM’). All subsequent analyses were performed using the detected ‘true’ CPM effect, i.e. the CPM timepoint which showed a significant ‘timepoint X paradigm’.

Secondly, it was analyzed whether this ‘true’ CPM effect was present in both cohorts in both areas. For that, 4 linear mixed models were performed (Supplementary Methods M3) (*n*-FDR: 2 per cohort).

#### Between-subject analyses

Group comparisons were performed for ‘true’ CPM effects in both areas (*n*-FDR: 2).

Additionally, participants were classified into CPM-inhibitors (PPT increase during CPM > 2 standard error of measurement (SEM) of PPTs), CPM-facilitators (PPT decrease > 2 SEM) or CPM-non-responders (PPT increase/decrease ≤ 2 SEM)^85^ (Supplementary Methods M5). Proportions of CPM-inhibitors, CPM-facilitators and CPM-non-responders in both areas were compared between the cohorts using Fisher’s exact tests (*n*-FDR: 2).

### Associations of Glx/GABA with CPM effects and experimental pressure pain sensitivity

Associations of Glx/GABA with ‘true’ CPM effects and experimental pressure pain sensitivity were tested using linear models because this allowed to test for ‘cohort’ (levels: ‘controls’, ‘CLBP’) interaction effects (’’true’ CPM effect X cohort’ and ‘experimental pressure pain sensitivity X cohort’), i.e. whether the associations differed between the cohorts. Associations were examined for both areas (*n*-FDR: 2 for ‘true’ CPM effects and 2 for experimental pressure pain sensitivity).

PPTs were used as a proxy for experimental pressure pain sensitivity. Here, PPTs from the first experimental session of the larger project were used to avoid a flawed analysis because a random string B-A (here the ‘true’ CPM effect) will typically correlate with a random string A (here PPTs before CPM). In the first experimental session, for each testing area, 3 PPTs were acquired.^71^ The average PPT of each area was used for analysis.

Two Spearman correlations were performed to explore whether the ‘true’ CPM effects depended on experimental pressure pain sensitivity in the controls, i.e. a ‘healthy’ state.

### Associations of Glx/GABA and CPM effects with clinical characteristics

In CLBP patients, Spearman correlations were used to investigate associations of Glx/GABA and ‘true’ CPM effects in both areas with clinical characteristics, i.e. (1) average clinical pain intensity over the past 4 weeks, (2) pain duration (in months), (3) spatial pain extent, and (4) within-MRS-session clinical pain or within-CPM-session clinical pain because ‘state’ pain can confound comparisons between pain and pain-free cohorts.^86^ (*n*-FDR: 4 for associations with Glx/GABA, *n*-FDR: 8 for associations with ‘true’ CPM effects). Using 3 additional Spearman correlations, associations of Glx/GABA with indicators of psychological distress in the CLBP patients, i.e. PCS and HADS anxiety and depression scores, were explored without multiple comparison correction due to the exploratory nature of this analysis.

### Confounding factors

The statistical analyses of proportional differences between CLBP patients and controls regarding menstrual cycle phases and pain-relevant medication intake, and associated potential influences on Glx/GABA or the parallel CPM effects are described in the Supplementary Methods M6.

### Data availability

Data of participants who gave informed consent for further use of their anonymized data will be made available upon request.

## Results

For all linear models and Welch’s t-tests, results with and without influential cases are presented in Supplementary Table 2.

### Participants

Out of 83 recruited participants (48 CLBP patients, 35 controls), 3 (CLBP patients) cancelled prior to the first session, 3 (2 CLBP patients, 1 control) were excluded due to a suspected neurological or psychiatric disorder and 1 (control) discontinued the scanning session due to discomfort. Five additional participants (2 CLBP patients, 3 controls) were excluded due to artefacts in the MR spectra (Supplementary Fig. 1). One additional control was excluded due to insufficient SNR.

The demographics of the final sample (41 CLBP patients, 29 controls) are described in Table 1.

**Table 1.** Participant demographics and clinical characteristics.

|  | **CLBP patients**<br>**(n = 41)** | **Controls**<br>**(n = 29)** | **Test**<br>**statistic** | **P** | **Effect size** |
| --- | --- | --- | --- | --- | --- |
| Age [years] | 54 (41 - 65) | 47 (34 - 67) | W = 553 | 0.625 | $r = 0.06$ |
| Sex (female:male) [n] | 22:19 | 17:12 |  | 0.810<sup>a</sup> |  |
| BMI [kg/m<sup>2</sup>] | 24.5 (3.47) | 23.3 (2.60) | $t = 1.5$ | 0.128 | $d = -0.37$ |
| PCS | 11 (5 - 22) | 3 (0 - 8) | W = 267.5 | **<0.001** | $r = 0.49$ |
| HADS anxiety | 4 (2 - 7) | 3 (1 - 5) | W = 485.5 | 0.193 | $r = 0.16$ |
| HADS depression | 1 (0 - 2) | 3 (1 - 6) | W = 314 | **<0.001** | $r = 0.41$ |
| Average clinical pain intensity over the past 4 weeks [NRS] | 4 (3 - 5) |  |  |  |  |
| Pain duration [months] | 106 (17 - 205.25)<sup>b</sup> |  |  |  |  |
| Spatial pain extent [%] | 1.3 (0.5 - 2.2) |  |  |  |  |
| Within-MRS-session clinical pain [NRS] | 1.5 (0 - 4) |  |  |  |  |
| Within-CPM-session clinical pain [NRS] | 2 (0 - 4) |  |  |  |  |

Values are presented as mean (SD) for continuous variables and as median (interquartile range) for ordinal or non-normally distributed variables. W-statistics refer to Wilcoxon rank-sum tests and t-statistics to unpaired t-tests.
<sup>a</sup>Fisher's exact test.
<sup>b</sup>N = 40 due to 1 missing value (participant did not indicate month of pain onset).

### Magnetic resonance spectroscopy

Shimming quality (FWHM H_2_O) and SNR indicated high spectral quality in both cohorts (Table 2, Fig. 2B and C). All metabolites were detected with adequate accuracy, i.e. mean Cramér-Rao lower-bounds (CRLB) below 20% (Supplementary Table 3).

**Table 2.** MRS outcomes, experimental pressure pain sensitivity and CPM effects.

|  | CLBP patients<br>(n = 41) | Controls<br>(n = 29) | n-IC | Test<br>statistic | P | Effect<br>size |
| --- | --- | --- | --- | --- | --- | --- |
| **MRS outcomes** |  |  |  |  |  |  |
| SNR | 19 (18 - 19) | 18 (17 - 21) |  | W = 567.5 | 0.748 | r = 0.04 |
| FWHM H<sub>2</sub>O [Hz] | 5.4 (0.91) | 5.4 (0.70) |  | t = 0.3 | 0.737 | d = 0.08 |
| FWHM NAA [Hz] | 4.3 (3.96 - 4.85) | 4.9 (3.96 - 4.85) |  | W = 545 | 0.561 | r = 0.07 |
| Glx/GABA | 4.0 (1.05) | 4.9 (1.17) | 1 | F = 10.7 | **0.002** | $\eta^2 = 0.14$ |
| Glx [mmol/kg] | 8.9 (1.34) | 9.9 (1.57) | 4 | F = 8.0 | **0.012<sup>a</sup>** | $\eta^2 = 0.11$ |
| GABA [mmol/kg] | 2.4 (0.67) | 2.1 (0.45) | 1<sup>†</sup> | F = 4.5 | **0.038<sup>a</sup>** | d = 0.46 |
| tCre [mmol/kg] | 6.5 (0.57) | 6.6 (0.46) | 4 | not meaningful<sup>b</sup> |  |  |
| tCho [mmol/kg] | 2.2 (0.24) | 2.2 (0.18) | 3 | not meaningful<sup>b</sup> |  |  |
| tmI [mmol/kg] | 8.9 (0.91) | 9.0 (1.03) | | F = 0.2 | 0.661 | $\eta^2 = 0.00$ |
| tNAA [mmol/kg] | 9.1 (1.16) | 9.0 (0.87) | | F = 0.2 | 0.654 | $\eta^2 = 0.00$ |
| GM [% of VOI] | 52.7 (5.19) | 50.2 (5.04) | 2<sup>†</sup> | F = 4.0 | 0.050 | $\eta^2 = 0.06$ |
| WM [% of VOI] | 41.2 (7.20) | 45.0 (6.0) | 2 | F = 5.4 | **0.024** | $\eta^2 = 0.07$ |
| CSF [% of VOI] | 6.0 (3.9) | 4.8 (2.5) | 3 | W = 510 | 0.312 | r = 0.12 |
| **Experimental pressure pain sensitivity** |  |  |  |  |  |  |
| PPT LB [kg/cm<sup>2</sup>] | 5.7 (2.76) | 6.1 (2.51) |  | t = -0.61 | 0.539 | d = 0.15 |
| PPT Hand [kg/cm<sup>2</sup>] | 4.0 (2.97 - 5.20) | 3.8 (2.83 - 4.90) |  | W = 553 | 0.625 | r = 0.06 |
| **CPM effects** |  |  |  |  |  |  |
|  | **CPM-SHAM**<br>**Controls**<br>**(n = 29)** | **CPM**<br>**Controls**<br>**(n = 29)** |  |  |  |  |
| PPT before Hand [kg/cm<sup>2</sup>] | 4.4 (1.75) | 4.2 (1.59) |  |  |  |  |
| PPT during Hand [kg/cm<sup>2</sup>] | 4.5 (1.49) | 5.1 (1.84) |  |  |  |  |
| PPT after Hand [kg/cm<sup>2</sup>] | 4.6 (1.63) | 4.6 (1.49) |  |  |  |  |
| ΔPPT parallel Hand [kg/cm<sup>2</sup>] | -0.02 (1.04) | -0.9 (1.21) | 9 | t<sup>ph</sup> = -2.8 | **0.010** |  |
| ΔPPT sequential Hand [kg/cm<sup>2</sup>] | -0.1 (0.96) | -0.4 (0.94) | 9 | t<sup>ph</sup> = -0.9 | 0.591 |  |
| ΔPPT parallel Hand [%] | -5.4 (25.15) | -27.6 (33.40) |  |  |  |  |
| ΔPPT sequential Hand [%] | -7.1 (26.99) | -14.7 (26.39) |  |  |  |  |
|  | **CLBP patients**<br>**(n = 41)** | **Controls**<br>**(n = 29)** |  |  |  |  |
| Cold-water bath LB<br>pain intensity [NRS] | 9 (7 - 10) | 8 (7 - 9) |  | W = 530 | 0.438 | r = 0.09 |
| Cold-water bath Hand<br>pain intensity [NRS] | 8 (6 - 9) | 8 (7 - 9) |  | W = 572 | 0.791 | r = 0.03 |
| PPT before LB [kg/cm<sup>2</sup>] | 5.6 (2.77) | 6.3 (2.38) |  |  |  |  |
| PPT during LB [kg/cm<sup>2</sup>] | 6.9 (2.96) | 8.0 (2.27) |  |  |  |  |
| CPM ΔPPT parallel LB [kg/cm<sup>2</sup>] | **-1.2 (1.37)\*\*\*** | **-1.6 (1.55)\*\*\*** |  |  |  |  |
| CPM ΔPPT parallel LB [%] | -25.8 (33.25) | -33.0 (34.15) | 4 | F = 0.75 | 0.388<sup>a</sup> | $\eta^2 = 0.01$ |
| PPT before Hand [kg/cm<sup>2</sup>] | 4.5 (2.09) | 4.2 (1.59) |  |  |  |  |
| PPT during Hand [kg/cm<sup>2</sup>] | 5.0 (2.0) | 5.1 (1.84) |  |  |  |  |
| CPM ΔPPT parallel Hand [kg/cm<sup>2</sup>] | **-0.5 (0.95)\*\*** | **-0.9 (1.21)\*\*\*** |  |  |  |  |
| CPM ΔPPT parallel Hand [%] | -16.9 (24.83) | -27.6 (33.40) | 3 | F = 2.3 | 0.274<sup>a</sup> | $\eta^2 = 0.03$ |
| CPM-inhibitors LB [%] | 47.5 | 53.5 |  |  |  |  |
| CPM-facilitators LB [%] | 7.5 | 3.5 |  |  | 0.794<sup>c</sup> |  |
| CPM-non-responders LB [%] | 45 | 43 |  |  |  |  |
| CPM-inhibitors Hand [%] | 33 | 56 |  |  |  |  |
| CPM-facilitators Hand [%] | 3 | 7 |  |  | 0.142<sup>c</sup> |  |
| CPM-non-responders Hand [%] | 64 | 37 |  |  |  |  |

Values are presented as mean (SD) for continuous variables and as median (interquartile range) for ordinal or not normally distributed variables. n-IC indicates the number of identified influential cases in the respective model. n-IC<sup>†</sup> indicates that removal of influential cases changed the statistical inference of the model. W-statistics refer to Wilcoxon rank-sum tests, t-statistics to unpaired t-tests or Sidak-corrected post-hoc tests (t<sup>ph</sup>) and F-statistics to linear models or Welch's test for GABA. $\eta^2$ values refer to partial $\eta^2$ s. Number of missing values for each variable are reported in Supplementary Table 6. LB: lower back.
<sup>a</sup>FDR-corrected for n = 2 tests.
<sup>b</sup>Due to the presence of a significant 'cohort X sex' interaction effect.
<sup>c</sup>Fisher's exact test with FDR-correction for n = 2 tests.
<sup>\*\*</sup>Statistically significant within-group CPM effect with P < 0.01, FDR-corrected for n = 2 tests
<sup>\*\*\*</sup>Statistically significant within-group CPM effect with P < 0.001, FDR-corrected for n = 2 tests

Glx/GABA was significantly lower in CLBP patients compared to controls (Table 2, Fig. 3A). This effect was driven by lower Glx as well as higher GABA in CLBP patients compared to controls (Table 2, Fig. 3B).

**Figure 3.**
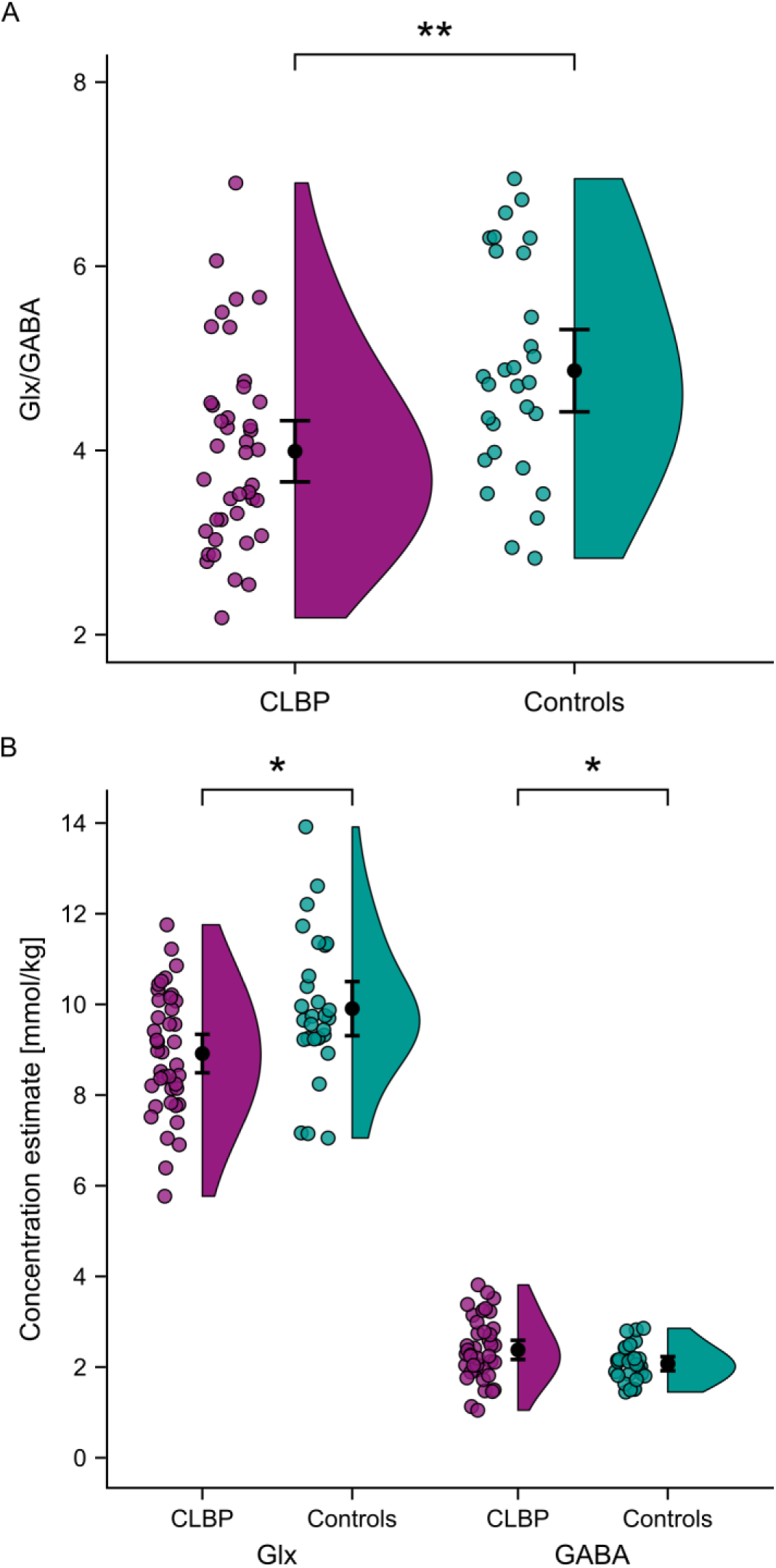
Lower Glx/GABA in CLBP patients driven by decreased Glx and increased GABA. The raincloud plots^144^ show the raw data (coloured dots), means and 95% confidence intervals (black dots and bars) and probability distributions (vertical “clouds”) of Glx/GABA (**A**) and Glx and GABA separately (**B**) for CLBP patients (purple) and controls (turquoise). \**P* <0.05, \*\**P* <0.01.

Sex had an influence on tCre and tCho in CLBP patients (significant ‘cohort X sex’ interactions; Supplementary Table 2) with highest concentrations observed in CLBP males. Additionally, tCre increased with age and tmI was higher in males (Supplementary Table 2). Otherwise, no metabolite differences were observed (Table 2). There were no differences in GM or CSF tissue fractions between the cohorts (Table 2). However, CSF tissue fractions increased with age in CLBP patients (*rho* = 0.73, *P* < 0.001) but not in controls (*rho* = 0.04, *P* = 0.831). Further, CLBP patients presented with lower WM tissue fractions compared to controls (Table 2).

Observed group differences in Glx or GABA were not driven by variations in GM or WM tissue fractions because within controls, neither Glx nor GABA correlated with GM (Glx: *r* = -0.23, *P* = 0.224; GABA: *r* = 0.04, *P* = 0.822) or WM tissue fractions (Glx: *r* = 0.32, *P* = 0.091, GABA: *r* = 0.10, *P* = 0.617).

Qualitatively similar cohort effects were observed using the literature-based water signals (Supplementary Table 4).

### Conditioned pain modulation

Cold water baths induced moderate to intense pain (Table 2). Six participants (5 CLBP, 1 control) reported low pain (NRS 1-3) in 1 or 2 cold water baths. Seven participants (5 CLBP, 2 controls) did not tolerate the full 2 min of 1 or 2 cold water baths. Two controls reported pain (NRS 1 and 2) during the ambient temperature water bath. For 1 CLBP patient and the respective matched control, CPM and experimental pressure pain sensitivity were assessed at the upper arm as control area because of scarring at the CLBP patient’s hand. These cases did not have an influence on the results.

First testing for the presence of a ‘true’ CPM effect beyond repeated-measures effects at the hand of the controls revealed a significant difference between the CPM and the CPM-SHAM effect on the PPTs (*F* = 4.2, *P* = 0.017). Inhibition of PPTs was stronger during CPM compared to during CPM-SHAM indicating a ‘true’ parallel CPM effect (Table 2, Fig. 4A). There was no difference in PPT changes after CPM compared to after CPM-SHAM (Table 2, Fig. 4A).

**Figure 4.**
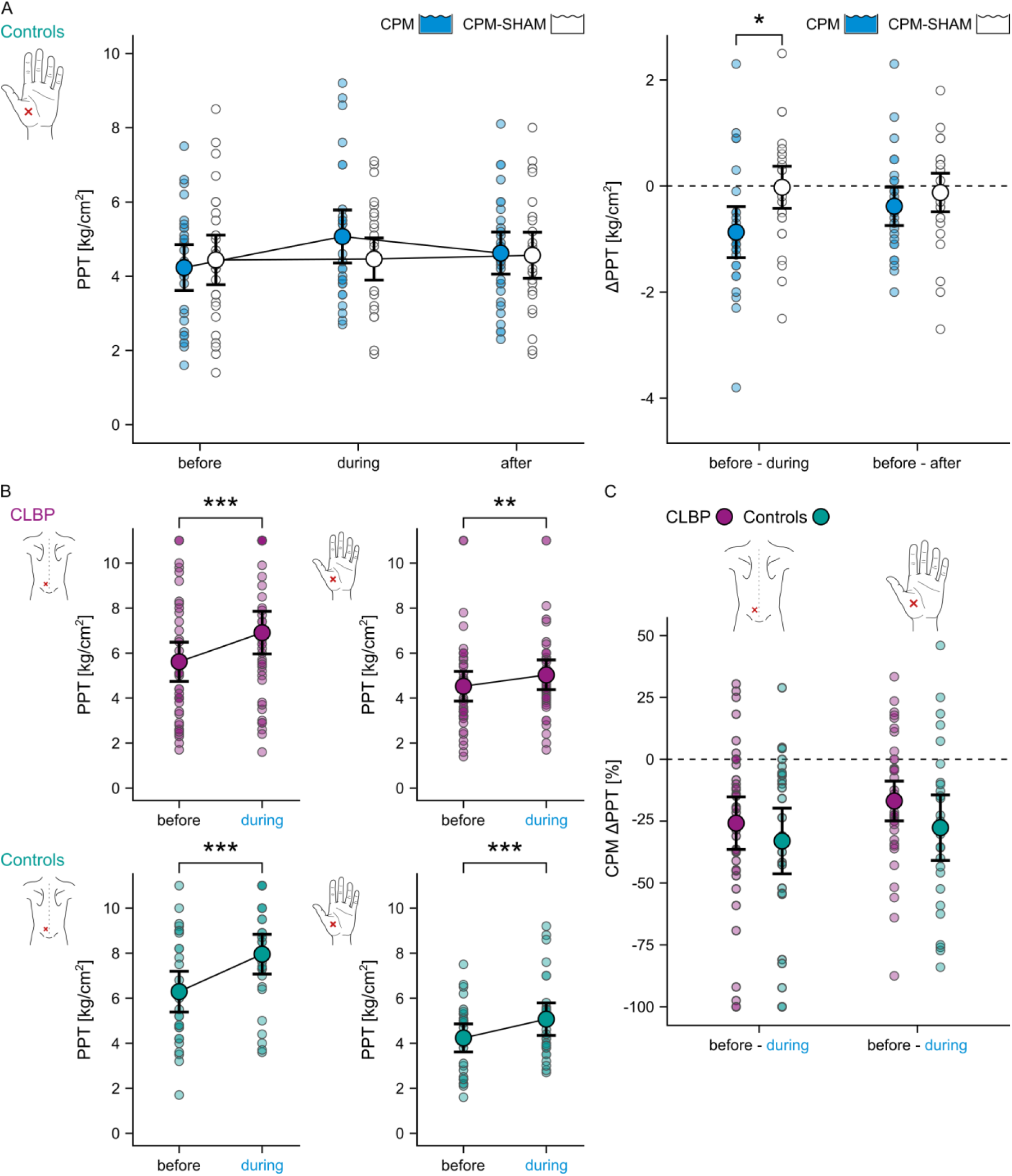
Inhibitory CPM effects on PPTs in CLBP patients and pain-free controls. (**A**) PPT changes in controls in response to the CPM (blue) and the CPM-SHAM (white) paradigm performed at the non-dominant hand (pain-free control area). Data are presented as PPT values at each timepoint of the 2 paradigms (left panel) and as absolute parallel (PPT before - PPT during) and sequential (PPT before - PPT after) PPT changes in response to the 2 paradigms (ΔPPT; right panel). (**B**) PPT values before and during the cold water bath reflecting parallel CPM effects at the lower back (most painful area) and at the non-dominant hand of CLBP patients (purple) and controls (turquoise). (**C**) Group comparison of relative parallel CPM effects (CPM ΔPPT), i.e. ([PPT before - PPT during] / PPT before) * 100, at the lower back and at the non-dominant hand. Negative absolute and relative ΔPPTs reflect inhibitory effects, positive values reflect facilitatory effects. The dotted line depicts a null effect. The plots represent the raw data (coloured semi-transparent dots), means (coloured dots) and 95% confidence intervals (black bars). \**P* <0.05, \*\**P* <0.01, \*\*\**P* <0.001.

This inhibitory parallel CPM effect on PPTs was significant in both cohorts at the lower back (CLBP: *F* = 31.4, *P* < 0.001, *n*-IC: 0, controls: *F* = 30.6, *P* < 0.001, *n*-IC: 1) and hand (CLBP: *F* = 11.2, *P* = 0.002, *n*-IC: 2, controls: *F* = 13.8, *P* < 0.001, *n*-IC: 2) (Table 2, Fig. 4B). Parallel CPM effects were not different between CLBP patients and controls (Table 2, Fig. 4C).

The SEM for PPTs was 12.4%. There was no difference in the proportions of CPM-inhibitors, CPM-facilitators and CPM-non-responders between the cohorts (Table 2).

### Associations of Glx/GABA with CPM effects and experimental pressure pain sensitivity

Associations of Glx/GABA with parallel CPM effects at the hand differed between the cohorts (’’true’ CPM effect X cohort’ interaction: *F* = 5.4, *P* = 0.046, *partial η^2^*= 0.08, n-IC: 1^†^). In controls, lower Glx/GABA was associated with larger CPM effects, but this association was not observed in CLBP patients (Fig. 5A). No cohort differences were observed for associations of Glx/GABA with parallel CPM effects at the lower back (Supplementary Table 2, Fig. 5A). Associations of Glx/GABA with PPTs differed between the cohorts at the lower back *and* hand (’experimental pressure pain sensitivity X cohort’ interaction: lower back: *F* = 9.0, *P* = 0.004, *partial η^2^* = 0.12, *n*-IC: 2; hand: *F* = 12.1, *P* = 0.002, *partial η^2^* = 0.16, *n*-IC: 2). Again, controls showed associations (lower Glx/GABA correlated with lower PPTs) that were not present in CLBP patients (Fig. 5B).

**Figure 5.**
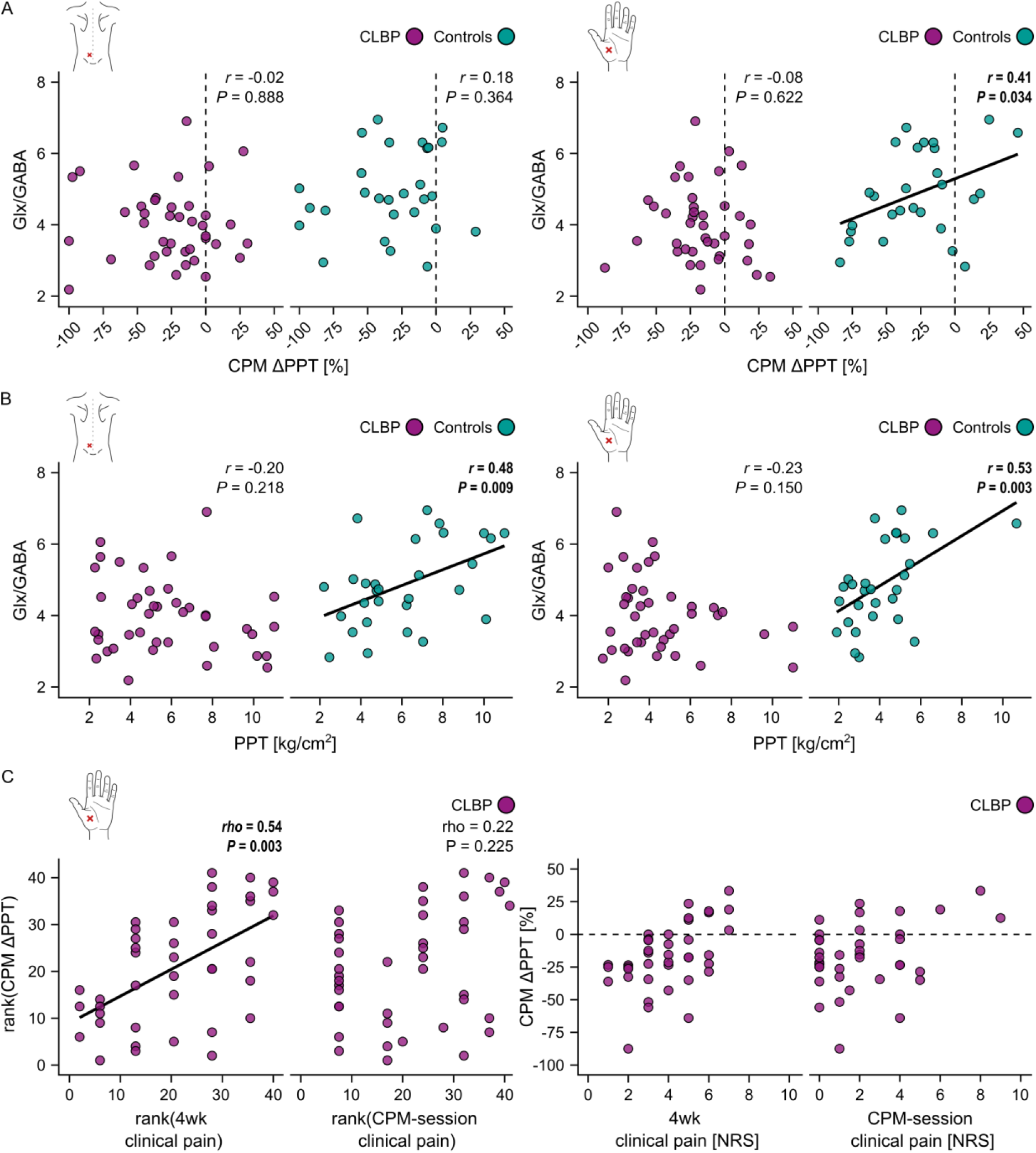
Glx/GABA- and CPM effect-related associations. (**A**) and (**B**) visualize the results of the linear models testing cohort differences in associations of Glx/GABA with ‘true’ CPM effects and experimental pressure pain sensitivity. Pearson correlations reflect the models’ interaction effects of interest, i.e. ‘’true’ CPM effect X cohort’ (significant for the non-dominant hand) and ‘experimental pressure pain sensitivity X cohort’ (significant for the lower back and the non-dominant hand). (**A**) shows Pearson correlations between Glx/GABA and relative parallel CPM effects (CPM ΔPPT) and (**B**) shows Pearson correlations between Glx/GABA and experimental pressure pain sensitivity, i.e. PPTs, of CLBP patients (purple) and controls (turquoise) at the lower back (most painful area) and at the non-dominant hand (pain-free control area). (**C**) Spearman correlations between relative parallel CPM effects (CPM ΔPPT) and average clinical pain intensities over the past 4 weeks (4wk clinical pain), i.e. ‘trait’ pain, and within-CPM-session clinical pain (CPM-session clinical pain), i.e. ‘state’ pain. Left panels show ranked data representative of Spearman correlations. To help with interpretability, the raw data is shown in the right panels. Negative relative CPM ΔPPTs reflect inhibitory CPM effects, positive relative CPM ΔPPTs reflect facilitatory CPM effects. The dotted line depicts a null CPM effect.

Parallel CPM effects depended on experimental pressure pain sensitivity, i.e. controls showed significantly smaller CPM effects with increasing PPTs, but exclusively at the hand (*rho* = 0.63, *P* < 0.001; lower back: *rho* = 0.22, *P* = 0.545).

### Associations of Glx/GABA and CPM effects with clinical characteristics of CLBP patients

Glx/GABA was not associated with average clinical pain intensities over the past 4 weeks, pain duration, spatial pain extent or within-MRS-session clinical pain (Supplementary Table 5). Smaller parallel CPM effects at the hand were observed for CLBP patients with higher average clinical pain intensities over the past 4 weeks (*rho* = 0.54, *P* = 0.003) but not with higher within-CPM-session clinical pain (*rho* = 0.22, *P* = 0.225) (Fig. 5C). No other associations between parallel CPM effects and clinical characteristics were observed (Supplementary Table 5). A statistical trend was observed for associations between Glx/GABA and PCS (*rho* = -0.29, *P* = 0.066) and HADS anxiety scores (*rho* = -0.29, *P* = 0.061). Glx/GABA was not related to HADS depression scores (*rho* = -0.19, *P* = 0.244).

### Confounding factors

Neither menstrual cycle phase nor pain medication intake had an influence on Glx/GABA or parallel CPM effects in either area (Supplementary Results R1).

## Discussion

This study presents evidence for PAG dysfunction in patients with CLBP. Firstly, CLBP patients showed a lower Glx/GABA ratio, i.e. a lower excitatory/inhibitory tone, in the PAG compared to pain-free controls, driven by decreased Glx *and* increased GABA. Secondly, while controls showed a link between Glx/GABA and experimental pressure pain sensitivity, these associations were disrupted in the patients. Additionally, CLBP patients with more severe clinical pain presented with reduced CPM capacities.

### Lower Glx/GABA in the PAG of CLBP patients

Growing evidence supports the presence of Glx and/or GABA imbalances across chronic pain conditions.^36^ For Glx and GABA, the investigated brain region matters regarding whether metabolite increases or decreases would be expected to contribute to enhanced pain sensitivity. In the PAG, a higher inhibitory or reduced excitatory tone might impede effective descending pain inhibition. This concept is supported by the observation of increased presynaptic GABA release and reduced glutamatergic neurotransmission in the PAG in animal models of chronic neuropathic pain.^40, 41^ The present study is the first to report similar changes in the PAG of patients with chronic pain. Intriguingly, the lower Glx/GABA in the PAG of CLBP patients compared to controls was due to a Glx decrease *and* a GABA increase. An earlier PAG ^1^H-MRS study reported Glx changes in the opposite direction in patients with episodic and chronic migraine.^45^ This discrepancy could theoretically be due to different pathomechanisms of migraine and CLBP, involving different sensory afferents, i.e. trigeminal or spinal afferents. However, both afferent types are similarly connected to the PAG, i.e. they terminate in the lateral PAG^87, 88^ and are inhibited upon PAG activation.^89–97^ Therefore, a methodological explanation is more plausible, for example the larger VOI size in the migraine study encompassing additional structures such as the red nucleus and parts of the substantia nigra.^45^ These regions show alterations in chronic migraine patients^98, 99^ and thus, might have contributed to the increased Glx in migraine patients.^45^ Other PAG ^1^H-MRS studies did not describe differences in Glx or GABA between patients and controls.^42–44^

The remaining metabolites did not show group differences except for the higher tCre and tCho in male CLBP patients. In addition, mI was increased in males compared to females. These findings are likely explained by the levels of all 3 metabolites increasing with age,^100, 101^ (as also observed in the present study for tCre) and higher levels in males,^102, 103^ because CLBP male patients were the oldest subgroup.

For ^1^H-MRS interpretation, consideration of brain tissue fractions within the VOI is crucial because metabolite concentrations are tissue-dependent.^104, 105^ Here, lower WM tissue fractions and higher CSF tissue fractions with increasing age were observed in CLBP patients. This pattern resembles age-related degenerative processes^106^ and might add to the notion of accelerated brain aging in chronic pain.^107, 108^ However, it can be ruled out that these factors drove the observed results in Glx or GABA because (1) CSF tissue fractions were accounted for in the metabolite concentration calculations, (2) lower WM tissue fractions would result in lower choline concentrations^109^ which was not observed in the CLBP patients, and (3) neither Glx nor GABA were related to WM tissue fractions in the controls. Lastly, Glx and GABA are predominantly present in GM^105, 110^ and even if WM tissue fractions had an influence, Glx and GABA would change in the same and not opposite direction.

OVERPRESS and voxel-based flip angle calibration minimize 2 limitations of conventional PRESS sequences, namely chemical shift displacement artifacts and the susceptibility to B_1_ inhomogeneities. A remaining limitation concerns reliable GABA detection because GABA’s signal overlaps with resonances from other metabolites. In the present study, adequate CRLBs of GABA were detected (absolute median: 58.9 I.U., relative median: 16.5 %) and the measured concentrations in controls were similar to expected concentrations in human brain tissue.^111, 112^ Nevertheless, the observed cohort difference in GABA could include influences from molecules with similar resonance frequencies to GABA. MEscher-GArwood PRESS^113^ sequences are more GABA-specific but unsuitable for the PAG because they require VOI sizes of approximately 30×30×30 mm^3114^ and are highly susceptible to frequency drifts.^115^

### Reduced CPM capacities in CLBP patients with more severe clinical pain

One strength of the present study is the use of a CPM-SHAM paradigm. This allowed for the detection of ‘true’ CPM effects^72, 85^ which were present for PPTs assessed in parallel to the cold water bath. The lack of a ‘true’ sequential CPM effect can be explained by sequential CPM effects being smaller compared to parallel CPM effects^116, 117^ due to decreasing CPM effects after cessation of the CS^118^ as well as distraction effects contributing to parallel CPM effects.^84^ Nevertheless, the present results support the combination of PPT as test stimulus and a cold water bath as CS as robust CPM paradigm.^69, 70^

The observed average ‘true’ parallel CPM effect magnitude of -30.3 % in the controls is consistent with existing reference values.^119, 120^ Impaired CPM effects were not observed in the CLBP patients, opposing the notion of deficient descending pain modulation in chronic pain conditions^121^ and some previous reports in CLBP.^122–124^ However, the recruited CLBP cohort presented with relatively low average clinical pain intensities over the past 4 weeks (median: NRS 4) and a meta-analysis indicated that CPM is only impaired in low back pain patients reporting high pain intensities (NRS >5).^125^ This is substantiated by the observed association between higher average clinical pain intensities and smaller CPM effects at the hand. Importantly, this association of CPM was only observed with ‘trait’ pain but not with the potentially confounding ‘state’ pain, indicating that CPM capacities relate to clinical pain characteristics and not to the sheer fact of being in pain during the CPM paradigm. The lack of an association between CPM effects at the lower back and average clinical pain intensities was potentially due to the extra-segmental application of the CS, reported to induce larger CPM effects compared to segmentally applied CS.^72, 126–128^ A similar pattern was observed here (lower back: CLBP: -25.8 %, controls: -33.0 %; hand: CLBP: -16.9 %, controls: -27.6 %). These overall larger CPM effects might have masked pain-related CPM variations at the lower back. Alternatively, CLBP patients may have a functional, protective descending pain inhibition of their lower back, but reduced capacities for the rest of their body.^129^

### Glx/GABA correlates with experimental pressure pain sensitivity in controls but not in CLBP patients

Contrary to our hypothesis, lower Glx/GABA was associated with larger CPM effects at the hand of the controls, i.e. more pain inhibition. However, the observed associations of controls with lower Glx/GABA being more sensitive to experimental pressure pain, i.e. showing lower PPTs, in both areas support our hypothesis of lower Glx/GABA being associated with less pain inhibition or pain facilitation. Most likely, this contradictory result has a methodological explanation, i.e. that exclusively at the hand, controls with higher PPTs showed smaller *relative* CPM effects probably for mathematical reasons (e.g. PPT increase by 3 kg/cm^2^ starting from 3 kg/cm^2^ = 100% increase vs. starting from 6 kg/cm^2^ = 50% increase) and due to the applied safety cut-off of 10 kg/cm^2^. At the lower back, where PPTs were higher compared to the hand and where no correlation between PPTs and CPM effects was present, Glx/GABA still correlated with the PPTs, but not with the CPM effects. Thus, Glx/GABA in the PAG might rather relate to experimental pressure pain sensitivity than to CPM effects.

DNIC are preserved after lesioning the PAG^130, 131^ and therefore, PAG function might not directly relate to CPM effects. Key roles for DNIC have been shown for medullary structures such as the subnucleus reticularis dorsalis^132, 133^ or the nucleus raphe magnus.^134^ Still, the PAG has modulatory effects on DNIC^135^ and is involved in RVM-driven descending (tonic) pain inhibition.^136^ Tonic descending pain inhibition has been shown to be particularly strong for deep tissue afferents.^137, 138^ Given this evidence, the present results might reflect a stronger tonic PAG-driven descending inhibition of deep tissue afferents in pain-free individuals with higher Glx/GABA.

CLBP patients did not show associations between Glx/GABA and experimental pressure pain sensitivity and thus, their tonic PAG-driven descending inhibition of deep tissue afferents might be dysregulated. Associations between PAG metabolites and descending pain inhibition have been suggested previously.^44^ PAG dysregulation might arise from altered inputs from supratentorial brain regions (reviewed in^4^). In chronic pain, a particularly relevant region with PAG connections is the amygdala.^4^ Preclinical pain models show alterations in amygdala-PAG pathways^139^ and increased amygdala activity resulting in decreased PAG activation via medial prefrontal cortex inhibition.^140–142^ Interestingly, human functional connectivity studies have shown altered connectivity patterns between these regions and the PAG in patients with chronic pain (schematic summary in Fig. 6). A possible association of Glx/GABA with amygdala-related factors is supported by the correlational trends between Glx/GABA and PCS or HADS anxiety scores in CLBP patients observed in the current study. Further research is needed to substantiate this hypothesis, for example studies integrating MRS, psychophysics and functional connectivity measures.

**Figure 6.**
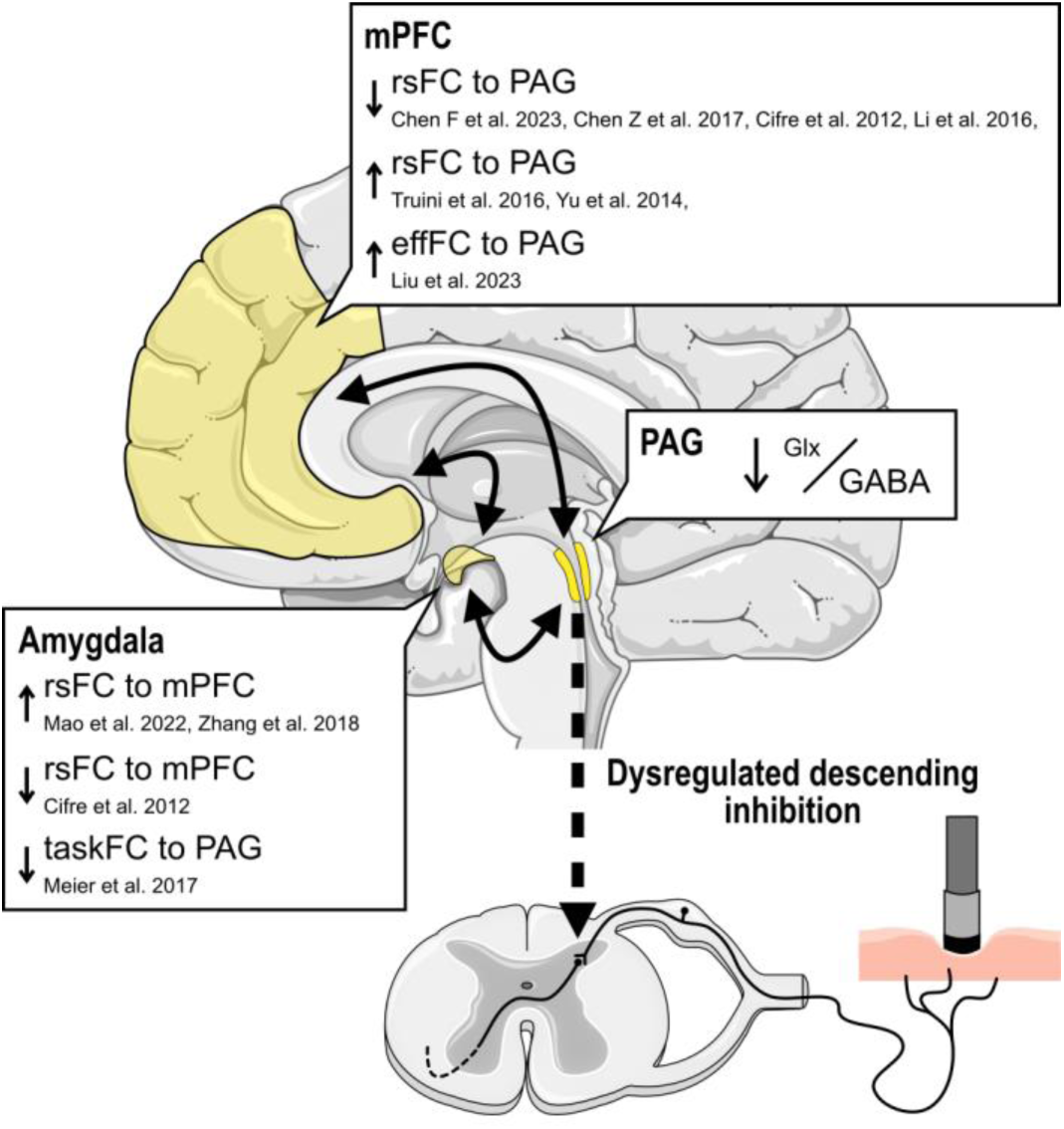
Clinical evidence for altered amygdala-medial prefrontal cortex (mPFC)-PAG connectivity patterns in chronic pain and potential consequences for PAG-driven descending pain inhibition. Summary of resting-state functional connectivity (rsFC) studies which found altered rsFC,^145–152^ altered effective rsFC (effFC)^153^ and altered task rsFC (during observation of harmful activities)^154^ in the amygdala-mPFC-PAG circuit in chronic pain patients compared to pain-free controls. Altered supratentorial input to the PAG might be associated with dysregulated PAG metabolites and PAG-driven descending inhibition, such as the decreased Glx/GABA and the lacking association between Glx/GABA and experimental pressure pain sensitivity in CLBP patients observed in the present study. Here, the anterior cingulate cortex was considered to be part of the mPFC.^155^ The schematic brain and spinal cord were adapted from Servier Medical Art (smart.servier.com).

### No further associations of Glx/GABA or CPM effects with clinical characteristics

Based on the present study, decreased Glx/GABA in CLBP appears to be independent of clinical pain intensity, pain duration and spatial pain extent. As discussed above, CPM effects were smaller in CLBP patients with more severe clinical pain. Previous reports documented reduced CPM capacities in CLBP patients with widespread pain.^143^ In the present study, no association between CPM effects and spatial pain extent was observed. This might be due to the relatively low spatial pain extent in the present CLBP cohort; using the widespread pain classification by Gerhardt and colleagues,^143^ only 1 of the included CLBP patients would have been classified as having widespread pain.

In summary, this study extends preclinical evidence by demonstrating a lower excitatory/inhibitory tone, i.e. lower Glx/GABA, in the PAG of patients with CLBP. Additionally, it provides insights into how the excitatory/inhibitory balance in the PAG relates to pain sensitivity. Lower Glx/GABA was related to higher experimental pressure pain sensitivity in pain-free controls while CLBP patients lacked this association, indicating a dysregulated PAG function. Independent of PAG metabolites, CLBP patients with more severe clinical pain showed reduced CPM capacities. It remains to be investigated whether these alterations are CLBP-specific, related to etiologies including deep afferent-associated mechanisms, such as musculoskeletal conditions, or generally applicable to chronic pain states.

## Acknowledgements

We thank all participants who took part in the study. Additionally, we thank Lucas Tauschek, Simon Carisch, Madeleine Hau and Alexandros Guekos for their support during data acquisition.

## Supplementary material

Supplementary material is available online.

## Abbreviations

CLBP: non-specific chronic low back pain
CPM: conditioned pain modulation
CRLB: Cramér-Rao lower bounds
CS: conditioning stimulus
DNIC: diffuse noxious inhibitory controls
FDR: false discovery rate
FWHM: full width at half maximum
Glx: glutamate + glutamine
GM: grey matter
^1^H-MRS: proton magnetic resonance spectroscopy
HADS: Hospital Anxiety and Depression Scale
MAD: median absolute deviation
NRS: numeric rating scale
PAG: periaqueductal grey
PCS: Pain Catastrophizing Scale
PPT: pressure pain threshold
PRESS: Point-RESolved Spectroscopy
RVM: rostral ventromedial medulla
SEM: standard error of measurement
SNR: signal-to-noise ratio
tCre: creatine + phosphocreatine
tCho: glycerophosphocholine + phosphocholine
tmI: myo-inositol + glycine)
tNAA: N-acetylaspartate + N-acetylaspartylglutamate
TE: echo time
TR: repetition time
TSP: temporal summation of pain
VOI: volume of interest
VSS: Very Selective Saturation
WM: white matter

