## Supplementary Material for "The periaqueductal grey in chronic low back pain: dysregulated metabolites and function"

#### Supplementary Methods

##### M1. Recruitment period, data collection period and sample size calculation

Participants were recruited between October 2019 and February 2022 (with a COVID-19 pandemic-related recruitment break from March to May 2020). Data were collected between December 2019 and April 2022.

To the best of our knowledge, at the time of protocol development, no study had examined Glx/GABA in the PAG and therefore, an a priori sample size calculation based on expected effect sizes was not possible. The sample size was thus chosen based on existing studies which showed differences in Glx or GABA between patients with different pain conditions and controls in other brain areas. Such MRS studies typically included around 19 participants per group.[1-20] Given the small size and the high level of physiological noise in the PAG, the aim was to recruit twice as many participants per group for the present study, i.e. 38 participants per group. In addition, due to an expected higher variability in the CLBP patients compared to pain-free controls, a 20 % larger sample size for the CLBP cohort, i.e. 46 CLBP patients, was aimed for. For a two-tailed independent t-test with a 5 % probability to commit a type I error ($\alpha = 0.05$), this sample size results in a power of 0.95 for a large effect (0.8), a power of 0.62 for a medium effect (0.5) and a power of 0.15 for a small effect (0.2) (calculated using G*Power 3.1.9.7[21]).

#### M2. Pain drawings for spatial pain extent assessment

Participants were instructed to shade their typically painful areas on printed standardized body charts (frontal and dorsal view). After manual contouring of the shaded areas, the pain drawings were digitalized and processed using a custom-made software that calculated the spatial extent of the shaded areas as a percentage of total body area. Only LBP-associated areas, i.e. the lower back, the buttocks and the legs, were considered for further analyses.

#### M3. Model specifications

##### $^{1}$H-MRS outcomes:

Linear models (R package 'stats' with the function 'lm') were performed with the respective $^{1}$H-MRS outcome as dependent variables and 'cohort' (levels: 'controls' and 'CLBP') as independent variable. For GABA, homogeneity of variance was not met and therefore, the 'cohort' effect was assessed using a Welch's t-test and age/sex influences were assessed in a separate linear model (for both cohorts combined because statistical inferences were not qualitatively different between the cohorts). For CSF tissue fractions, homogeneity of variance was not met and data was not normally distributed and therefore, the 'cohort' effect and sex influences were assessed using Wilcoxon rank-sum tests and age influences were tested using a Spearman correlation.

##### CPM effects:

*Within-subject analyses:* The linear mixed model (R package 'nlme', function 'lmer') assessing the presence of 'true' CPM effects beyond repeated-measures was performed on the data of the controls with PPT as dependent variable and 'timepoint' (levels: 'before', 'during', and 'after') and 'paradigm' (levels: 'CPM', 'CPM-SHAM') and the interaction of interest 'timepoint X paradigm' as independent variables and the participants' identifier as random effect. Post-hoc tests were performed using planned comparisons ('before-during X CPM-CPM-SHAM' and 'before-after X CPM-CPM-SHAM'; R functions 'emmeans' followed by 'contrast') with Sidak's *P*-value adjustment. Age/sex influences were not assessed in this model because age/sex-dependent differences between the CPM and CPM-SHAM paradigm were not expected and because age/sex influences on CPM effects were tested in the models described below.

The 4 linear mixed models assessing 'true' CPM effects in both cohorts in both areas were performed with PPT as dependent variable and 'timepoint' (levels: 'before', 'during') as independent variable and the participants' identifier as random effect.

*Between-subject analyses:* The linear model assessing group differences in 'true' CPM effects in both areas were performed with the 'true' CPM effect as dependent variable and 'cohort' as independent variable. Only age/sex main effects were tested because age/sex interactions with cohort allocation had already been tested in the within-subject analyses.

**Associations of Glx/GABA with CPM effects and experimental pressure pain sensitivity:**

The linear models were performed with Glx/GABA as dependent variable and the 'true' CPM effect or the PPTs (proxy for experimental pressure pain sensitivity), 'cohort' and the interaction of interest "true' CPM effect X cohort' or 'PPT X cohort' as independent variables.

#### M4. Statistical analysis of age/sex influences and assessment of influential cases

For all linear models, potential influences of age and sex on the dependent variables were examined by analyzing age and sex main effects and interaction effects with the model's independent variables. The models were always fitted with all interactions first, except for 'age X sex' interactions, which were not tested because the number of participants of different ages within a certain sex was low and this interaction was not of interest here. If the age/sex interactions were not significant, they were removed from the model to assess age/sex main effects. If the age/sex main effects were not significant, they were also removed from the model. Of note, the effects of interest were always left in the model when assessing age/sex influences.

Influential cases are outliers in the model residuals which exert a large influence over the model.[22] Influential cases were identified based on standardized residuals and Cook's distance.[22] For standardized residuals, a cut-off of 1.96 was chosen.[22] For Cook's distance, a conservative cut-off of 4/(n-k-1)[23] (with n = number of observations and k = number of predictors in the model) was chosen to minimize false-positive or false-negative results driven by a small number of observations. For GABA, where the group comparison was made using a Welch's t-test, outliers > ±2.5 MAD were treated as influential cases.

#### M5. Standard error of measurement of pressure pain thresholds calculation

The standard error of measurement (SEM) for PPTs was calculated using data of all controls from the larger project which were measured at the hand as control area and randomized to perform the CPM-SHAM paradigm first (N=30) with the formula:[24]

$$SEM\ [\%] = \frac{SD(PPT\ before\ CPM - SHAM)\ x\ \sqrt{1 - ICC}}{mean(PPT\ before\ CPM - SHAM)}\ x\ 100$$

The ICC (two-way, absolute agreement, single rater/measurement) was calculated between the PPT before and the PPT during CPM-SHAM. Participants were classified as CPM-inhibitors or CPM-facilitators if they presented with a PPT increase or PPT decrease, respectively, during CPM >2 SEM and otherwise as CPM-non-responders.[24]

#### M6. Statistical analysis of confounding factors

Proportions of the different menstrual cycle phases (menstruation, follicular, ovulation, or luteal) and of participants reporting regular pain-relevant medication intake were compared between the CLBP patients and controls using Fisher's exact tests.

Because the proportions of participants in the different menstrual cycle phases (for 22 CLBPs and 17 controls: *n* without menstrual cycle: 14 CLBP, 11 controls – MRS session: *n* menstruation: 2 CLBP, 2 controls; *n* follicular: 2 CLBP, 0 controls; *n* luteal: 3 CLBP, 3 controls; *n* ovulation: 1 CLBP, 1 controls – CPM session: *n* menstruation: 3 CLBP, 1 controls; *n* follicular: 4 CLBP, 3 controls; *n* luteal: 1 CLBP, 2 controls) were not different between the 2 cohorts (Fisher's exact test: MRS session: $P = 0.783$, CPM session: $P = 0.885$), data of the 2 cohorts were pooled to assess whether the menstrual cycle phase had an influence on Glx/GABA or the parallel CPM effects in both areas using Kruskal-Wallis tests (without multiple comparison correction to minimize the risk for false negatives).

Similarly, because the proportions of participants with and without pain-relevant medication intake (8/33 CLBP, 2/27 controls) were not different between the 2 cohorts (Fisher's exact test: $P$ = 0.178), data of the 2 cohorts were pooled to assess whether regular pain-relevant medication intake had an influence on Glx/GABA or the parallel CPM effects in both areas using Wilcoxon rank-sum tests (without multiple comparison correction to minimize the risk for false negatives).

### Supplementary Results

#### R1. Confounding factors

Glx/GABA was not different between participants measured in different menstrual cycle phases ($\chi^2$ = 1.9, $P$ = 0.586), nor between participants with or without pain-relevant medication intake ($W$ = 409, $P$ = 0.069).

Also, the parallel CPM effects in both areas were not different between participants measured in different menstrual cycle phases (hand: $\chi^2$ = 7.0, $P$ = 0.137; lower back: $\chi^2$ = 1.1, $P$ = 0.895), nor between participants with or without pain-relevant medication intake (hand: $W$ = 244, $P$ = 0.823; lower back: $W$ = 276, $P$ = 0.815).

#### Supplementary Figures

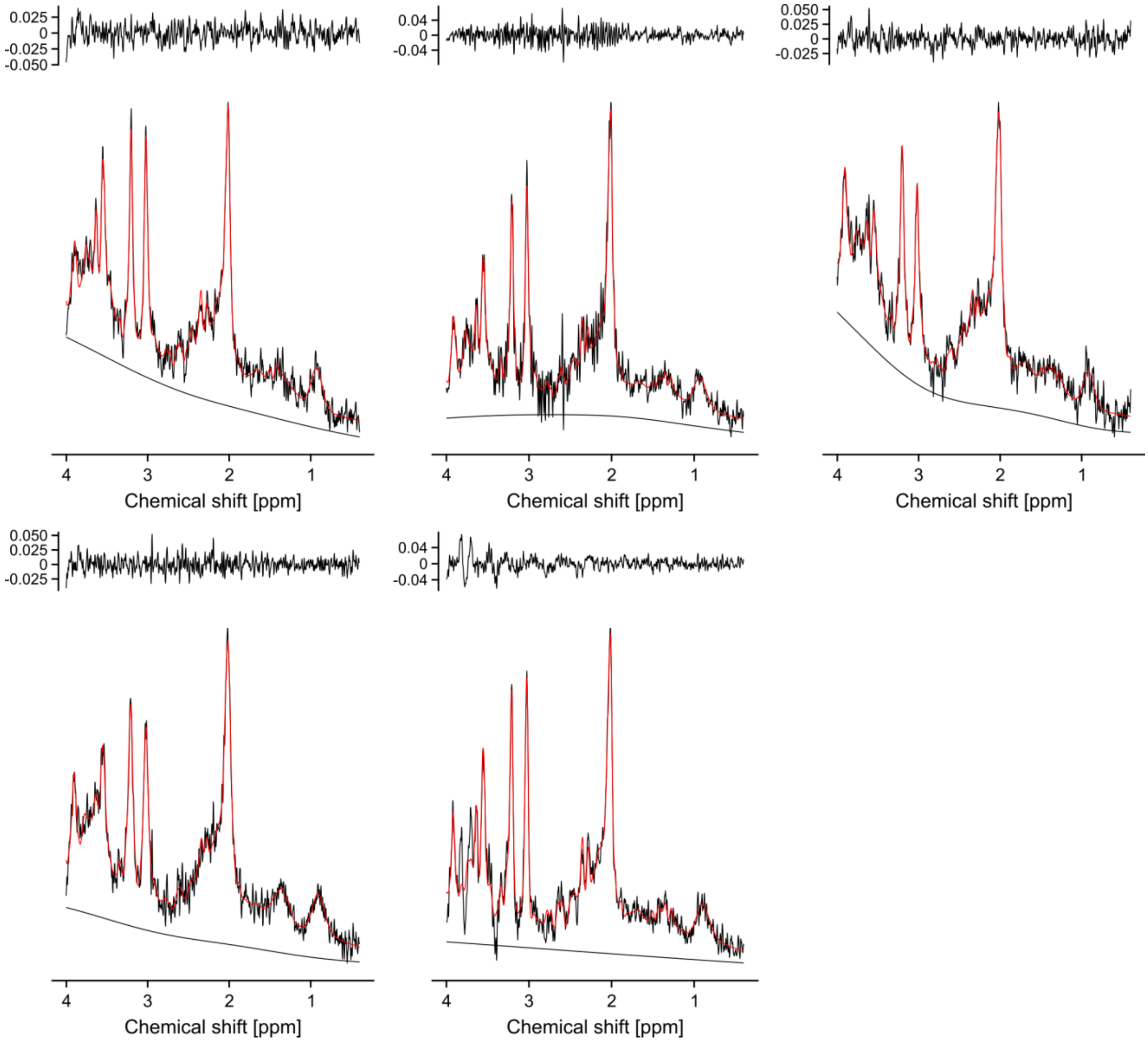


**Supplementary Figure 1 Excluded MR spectra due to the presence of artefacts.** Spectra were excluded from 2 CLBP patients and 3 controls.

### Supplementary Tables

**Supplementary Table 1 Technical details of the $^{1}$H-MRS acquisition using the experts' consensus checklist for a single voxel $^{1}$H-MRS study.[25]**

| | |
|---|---|
| ***1. Hardware*** | |
| a. Field strength [T] | 3 T |
| b. Manufacturer | Philips |
| c. Model (software version if available) | Achieva with dStream Upgrade. Software Release 5.6.1. |
| d. RF coils: nuclei (transmit/receive), number of channels, type, body part | 32-channel dStream receive-only phased-array head coil |
| e. Additional hardware | N/A |
| ***2. Acquisition*** | |
| a. Pulse sequence | PRESS in combination with 6 saturation pulses (OVERPRESS)[26-28] and a voxel-based flip angle calibration performed prior to each measure-ment to achieve the desired flip angle and thus optimal SNR.[29,30] |
| b. Volume of Interest (VOI) locations | Periaqueductal grey |
| c. Nominal VOI size [cm3, mm3] | 11x15x18 $mm^3$ (APxRLxFH). Accounting for the saturation pulses, the resulting effective VOI size was 8.8x10.2x12.2 $mm^3$=1.1 mL. |
| d. Repetition Time (TR), Echo Time (TE) [ms, s] | TR: 2500ms, TE: 33ms |
| e. Total number of Excitations or acquisitions per spectrum | 512 acquisitions per spectrum divided into 8 blocks of 64 averages each. At the beginning of each block, 1 scan without water suppression was performed, resulting in a total of 8 acquisitions for the water reference (WR-shortTR).<br><br>Further, after completion of the 8 blocks, an additional water reference scan with TR 10000ms was performed within the same VOI (WR-longTR). TEs were varied for the 6 acquired averages (+ 2 dummy scans), i.e. 33/66/107/165/261/600 ms, allowing to estimate the $T_2$ relaxation time of water within the VOI and therewith, obtain a subject-specific approxi-mation of the fully-relaxed water signal within the VOI.[31] All settings were kept identical to the previous sequence except the center frequency of the applied pulses which was set to the resonance frequency of water instead of creatine. |
| f. Additional sequence parameters (spectral width in Hz, number of spectral points, frequency offsets) | Spectral width 2000 Hz, 2048 points. |
| g. Water Suppression Method | VAriable Power radiofrequency pulses with Optimized Relaxation delays (VAPOR) |
| h. Shimming Method, reference peak, and thresholds for "acceptance of shim" chosen | Second-order automatic pencil-beam shim |
| ***3. Data analysis methods and outputs*** | |
| a. Analysis software | ReconFrame (GyroTools LLC, Zurich, Switzerland) to pre-process the spectra and LCModel version 6.3[32] for analysis. For the pre-processing, code from FID-A[33] was added to ReconFrame. |
| b. Processing steps deviating from quoted reference or product | Processing of .raw/.lab with ReconFrame including a) eddy current correction[34] and b) coil combination.<br><br>Then, for the processing with spectral registration: Frequency alignment using spectral registration in the time domain[35] (adopted from FID-A[33]). For the spectral registration in the time domain, data was 8-fold zero-filled and filtered with a 2 Hz Gaussian filter. Only the first 500 ms were used for alignment and the single averages were aligned to the median of all averages.<br><br>Without spectral registration:<br>Minimal frequency alignment was achieved by the performed eddy current correction with the interleaved water unsuppressed scans (WR-shortTR scan).<br><br>Both approaches were followed by the following steps: c) residual water filtering, d) 1Hz Gaussian filtering, and e) FWHM of water peak (FWHM H2O; second unsuppressed water peak from WR-shortTR) estimation (adopted from FID-A[33]). |
| c. Output measure<br><br>(e.g. absolute concentration, institutional units, ratio) | Ratio to water signal from WR-shortTR or WR-longTR. Ratios were corrected for CSF fraction and multiplied with the inverse of the molecular weight of water. Relaxation attenuation of the metabolite signals was not corrected. With that, a rough estimate |

| | |
|---|---|
| | of moles of metabolite per mass of tissue water (excluding CSF) - molar concentration mol/kg, was achieved.<br><br>Based on the different WR scans, the fully relaxed water signal was estimated differently:<br><br>WR-shortTR:<br>The WR-shortTR water scan was provided as water reference to LCModel. Relaxation attenuation of the water signal was considered based on literature values. The following $T_1$ /$T_2$ values were used for the different tissue types (ms): GM: 1820 / 100; WM: 1080 / 70; CSF: 4160 / 500 and the following relative densities of Nuclear Magnetic Resonance-visible water: GM: 0.78, WM: 0.65, CSF: 0.97.<br><br>WR-longTR:<br>In this case the WR-longTR water scan was provided as water reference to LCModel. The fully-relaxed water signal was estimated based on the measured subject-specific TE series. The decay of the water was fitted with an exponential decay within MATLAB 2022 using "fitnlm" and used to estimate the water peak area at TE=0 ms. The ratio of the water peak area at TE=33 ms and TE=0 ms was used to correct the conc. values resulting from LCModel.<br><br>General:<br>Relative tissue type volume fractions within the VOI were determined using the $T_1$-weighted planning images (three-dimensional magnetization-prepared rapid gradient-echo (MPRAGE) sequence,[36] 1mm$^3$ isotropic, TE=3.7 ms, TR=8.1 ms, TI=1024 ms, shot interval=3000 ms, field of view: 240x160x240 mm$^3$ (APxLRxFH), flip angle=8°, scan time=7 min32 s) which were segmented using SPM12.[37]<br><br>For both approaches, based on WR-shortTR and based on WR-longTR, ratios to water signal were obtained from LCModel with WCONC=55556 and ATTH2O=1. |
| d. Quantification references and assumptions, fitting model assumptions | The unsuppressed water peak (WR-shortTR or WR-longTR) was used as reference.<br><br>A simulated basis set containing the following 20 metabolites was used to determine peak areas in the chemical shift range from 0.4 ppm and 4.0 ppm:<br>alanine, aspartate, glucose, creatine, phosphor-creatine, glutamine, glutamate, glycerol-phosphocholine, phosphocholine, lactate, myo-inositol, N-acetylaspartate, N-acetylaspartyl-glutamate, scyllo-inositol, glutathione, taurine, glycine, phosphoethanolamine, ascorbate, and γ-aminobutyric acid.<br><br>Basis set simulations were performed using FID-A[33] (https://github.com/CIC-methods/FID-A retrieved commit from 2022 05 01), an open-source software toolkit for the simulation and processing of MRS data. 2D simulations were carried out over the effective voxel size (assuming ideal saturation bands) with a spatial resolution of 40 × 40 points in the directions of the refocusing pulses and using the actual pulse shape of the refocusing pulses.<br><br>Simulated contribution of macromolecules and lipid signals were provided within LCModel. |
| ***3. Data quality*** | |
| a. Reported variables<br><br>(SNR, Linewidth (with reference peaks)) | SNR and FWHM of the N-acetylaspartate peak obtained from the LCModel output. To assess the shim quality in the VOI, the FWHM of the water peak (FWHM $H_2O$) from the water reference scan (WR-shortTR) was determined. |
| b. Data exclusion criteria | Visual inspection of artifacts and spectra with FWHM $H_2O$ values above 2.5 mean absolute deviance (MAD)[38] of the group median or SNR values below 2.5 MAD of the group median. |
| c. Quality measures of postprocessing Model fitting (e.g. CRLB, goodness of fit, SD of residual) | Absolute CRLBs[39] of selected metabolites, i.e. relative % CRLBs obtained from LCModel multiplied by the conc. values obtained from LCModel. |
| d. Sample Spectrum | Figure 2 |

**Supplementary Table 2 Linear model and Welch's test results with and without influential cases.**

| Dependent variable | | *n*-IC | Independent variables | | | | | | | | | | | |
|---|---|---|---|---|---|---|---|---|---|---|---|---|---|---|
| **MRS outcomes** | | | | | | | | | | | | | | |
| | **Influential cases** | | *F* | *P* | *F* | *P* | *F* | *P* | *F* | *P* | *F* | *P* | *F* | *P* |
| | | | **cohort** | | **age** | | **sex** | | **cohort X age** | | **cohort X sex** | | | |
| Glx/GABA | with | 1 | 10.7 | **0.002** | 2.6 | 0.114 | 0.6 | 0.442 | 0.0 | 0.965 | 2.6 | 0.115 | | |
| | without | | 13.6 | **<0.001** | | | | | | | | | | |
| Glx [mmol/kg] | with | 4 | 8.0 | **0.012**[a] | 0.7 | 0.634[a] | 2.9 | 0.155[a] | 1.2 | 0.287 | 0.5 | 0.488 | | |
| | without | | 9.1 | **0.007**[a] | | | | | | | | | | |
| GABA[b] [mmol/kg] | with | 1[†] | 3.8 | 0.055[a] | 0.2 | 0.634[a] | 2.1 | 0.155[a] | | | | | | |
| | without | | 4.5 | **0.038**[a] | | | | | | | | | | |
| tCre [mmol/kg] | with | 4 | not meaningful[c] | | 8.2 | **0.006** | not meaningful[c] | | 0.4 | 0.525 | 6.5 | **0.013** | | |
| | without | | not meaningful[c] | | 9.5 | **0.003** | not meaningful[c] | | | | 17.5 | **<0.001** | | |
| tCho [mmol/kg] | with | 3 | not meaningful[c] | | 0.2 | 0.619 | not meaningful[c] | | 0.4 | 0.513 | 6.5 | **0.016** | | |
| | without | | not meaningful[c] | | | | not meaningful[c] | | | | 6.0 | **0.017** | | |
| tmI [mmol/kg] | with | 4 | 0.2 | 0.661 | 1.6 | 0.209 | 9.8 | **0.003** | 0.0 | 0.958 | 3.4 | 0.068 | | |
| | without | | 0.2 | 0.644 | | | 16.5 | **<0.001** | | | | | | |
| tNAA [mmol/kg] | with | 4 | 0.2 | 0.654 | 2.7 | 0.107 | 0.4 | 0.529 | 3.3 | 0.075 | 2.2 | 0.141 | | |
| | without | | 0.0 | 0.949 | | | | | | | | | | |
| GM [% of VOI] | with | 2[†] | 4.0 | **0.049** | 0.0 | 0.984 | 0.3 | 0.611 | 0.2 | 0.674 | 0.0 | 0.903 | | |
| | without | | 4.0 | 0.050 | | | | | | | | | | |
| WM [% of VOI] | with | 2 | 5.4 | **0.024** | 2.7 | 0.104 | 2.7 | 0.105 | 1.2 | 0.273 | 0.2 | 0.695 | | |
| | without | | 6.5 | **0.013** | | | | | | | | | | |
| CSF[d] [% of VOI] | | | *W* = 510 | 0.312 | CLBP:<br>*rho* = 0.73<br>HC:<br>*rho* = 0.04 | CLBP:<br>**<0.001**<br>HC:<br>0.831 | CLBP:<br>*W* = 136<br>HC:<br>*W* = 83 | CLBP:<br>0.056<br>HC:<br>0.405 | | | | | | |
| **CPM effects** | | | | | | | | | | | | | | |
| | | | **timepoint** | | **paradigm** | | **timepoint X paradigm** | | | | | | | |
| PPT Hand (controls) [kg/cm$^2$] | with | 8 | not meaningful[c] | | not meaningful[c] | | 4.2 | **0.017** | | | | | | |
| | without | | not meaningful[c] | | not meaningful[c] | | 4.7 | **0.011** | | | | | | |
| | | | **timepoint** | | **timepoint X age** | | **timepoint X sex** | | | | | | | |
| PPT LB (CLBP) [kg/cm$^2$] | with | 0 | 31.4 | **<0.001**[a] | 3.5 | 0.142[a] | 0.1 | 0.992[a] | | | | | | |
| | without | | | | | | | | | | | | | |
| PPT Hand (CLBP) [kg/cm$^2$] | with | 2 | 11.2 | **0.002**[a] | 0.1 | 0.736[a] | 0.0 | 0.992[a] | | | | | | |
| | without | | 20.1 | **<0.001**[a] | | | | | | | | | | |

| | | | | | | | | | | | | | | |
|---|---|---|---|---|---|---|---|---|---|---|---|---|---|---|
| PPT LB (controls) [kg/cm$^2$] | with | 1 | 30.6 | **<0.001[a]** | 4.8 | 0.076[a] | 0.1 | 0.710[a] | | | | | | |
| | without | | 47.1 | **<0.001[a]** | | | | | | | | | | |
| PPT Hand (controls) [kg/cm$^2$] | with | 2 | 13.8 | **<0.001[a]** | 2.6 | 0.119[a] | 0.6 | 0.710[a] | | | | | | |
| | without | | 23.1 | **<0.001[a]** | | | | | | | | | | |
| | | | **cohort** | | **age** | | **sex** | | | | | | | |
| CPM ΔPPT parallel LB [%] | with | 4 | 0.8 | 0.388 [a] | 4.3 | 0.064[a] | 2.6 | 0.217[a] | | | | | | |
| | without | | 0.6 | 0.429[a] | | | | | | | | | | |
| CPM ΔPPT parallel Hand [%] | with | 3 | 2.3 | 0.274 [a] | 3.6 | 0.064[a] | 0.2 | 0.670[a] | | | | | | |
| | without | | 4.2 | 0.088[a] | | | | | | | | | | |
| **Associations of Glx/GABA with CPM effects** | | | | | | | | | | | | | | |
| **Lower back:** | | | **CPM ΔPPT parallel** | | **CPM ΔPPT parallel X cohort** | | **CPM ΔPPT parallel X age** | | **CPM ΔPPT parallel X sex** | | **CPM ΔPPT parallel X cohort X age** | | **CPM ΔPPT parallel X cohort X sex** | |
| Glx/GABA | with | | 0.3 | 0.596[a] | 0.7 | 0.409[a] | 2.1 | 0.315[a] | 2.1 | 0.197[a] | 0.4 | 0.553[a] | 0.5 | 0.942[a] |
| | without | | 0.1 | 0.707 | 2.2 | 0.147[a] | | | | | | | | |
| **Hand:** | | | **CPM ΔPPT parallel** | | **CPM ΔPPT parallel X cohort** | | **CPM ΔPPT parallel X age** | | **CPM ΔPPT parallel X sex** | | **CPM ΔPPT parallel X cohort X age** | | **CPM ΔPPT parallel X cohort X sex** | |
| Glx/GABA | with | 1 | 1.9 | 0.354[a] | 3.6 | 0.122[a] | 0.3 | 0.598[a] | 1.7 | 0.197[a] | 0.5 | 0.553[a] | 0.0 | 0.942[a] |
| | without | | not meaningful[c] | | 5.4 | **0.046[a]** | | | | | | | | |
| **Associations of Glx/GABA with experimental pressure pain sensitivity** | | | | | | | | | | | | | | |
| **Lower back:** | | | **PPT** | | **PPT X cohort** | | **PPT X age** | | **PPT X sex** | | **PPT X cohort X age** | | **PPT X cohort X sex** | |
| Glx/GABA | with | 2 | not meaningful[c] | | 9.0 | **0.004[a]** | 0.6 | 0.628[a] | 0.1 | 0.786[a] | 0.8 | 0.481[a] | 1.6 | 0.408[a] |
| | without | | not meaningful[c] | | 15.1 | **<0.001[a]** | | | | | | | | |
| **Hand:** | | | **PPT** | | **PPT X cohort** | | **PPT X age** | | **PPT X sex** | | **PPT X cohort X age** | | **PPT X cohort X sex** | |
| Glx/GABA | with | 2 | not meaningful[c] | | 12.1 | **0.002[a]** | 0.2 | 0.628[a] | 0.1 | 0.786[a] | 0.5 | 0.481[a] | 0.5 | 0.468[a] |
| | without | | not meaningful[c] | | 14.7 | **<0.001[a]** | | | | | | | | |

LB: lower back.
[a]FDR-corrected for $n = 2$ tests.
[b]Welch's test followed by linear model for age/sex influence assessment.
[c]Due to the presence of a significant interaction effect.
[d]Wilcoxon tests for cohort and sex differences and Spearman correlation for age influence assessment.
*n*-IC[†]: statistical inference changed with removal of influential cases.

**Supplementary Table 3 CRLBs of measured metabolites using MRS.**

| | CLBP patients (*n* = 41) | Controls (*n* = 29) |
|---|---|---|
| **Absolute CRLBs[a]** | | |
| Glx [I.U.] | 82.9 (76.69 - 93.91) | 85.2 (79.74 - 93.69) |
| GABA [I.U.] | 58.5 (54.46 - 63.91) | 59.3 (53.62 - 66.94) |
| tCre [I.U.] | 21.1 (20.37 - 22.42) | 22.3 (21.60 - 23.15) |
| tCho [I.U.] | 7.8 (7.10 - 9.97) | 7.9 (7.35 - 10.12) |
| tmI [I.U.] | 39.4 (30.73 - 43.78) | 37.6 (32.08 - 42.37) |
| tNAA [I.U] | 29.3 (28.07 - 31.06) | 29.4 (28.00 - 30.56) |
| **Relative CRLBs** | | |
| Glx [%] | 6 (5 - 7) | 5 (5 - 6) |
| GABA [%] | 15 (14 - 19) | 18 (15 - 20) |
| tCre [%] | 2 (2 - 2) | 2 (2 - 2) |
| tCho [%] | 2 (2 - 3) | 2 (2 - 3) |
| tmI [%] | 3 (2 - 3) | 3 (2 -3) |
| tNAA [%] | 2 (2 - 2) | 2 (2- 2) |

Because not all outcome measures were normally distributed and to allow comparison between the CRLBs of different metabolites, all values are reported as median (interquartile range).
[a]Calculated by multiplying the relative CRLBs with the metabolite concentration as ratio to water from the LCModel output.

**Supplementary Table 4 MRS outcomes using literature-based water signals for metabolite quantification.**

| | *n*-IC | CLBP patients ($n$ = 41) | Controls ($n$ = 29) | Test statistic | *P* | Effect size |
|---|---|---|---|---|---|---|
| **MRS outcomes** | | | | | | |
| Glx/GABA | 1 | 4.0 (1.05) | 4.9 (1.17) | $F$ = 10.7 | **0.002** | $\eta^2$ = 0.14 |
| Glx [mmol/kg] | 4 | 8.3 (1.14) | 9.3 (1.43) | $F$ = 10.0 | **0.005**[a] | $\eta^2$ = 0.13 |
| GABA [mmol/kg] | 2† | 2.2 (0.62) | 2.0 (0.43) | $F$ = 4.3 | **0.043**[a] | $d$ = 0.44 |
| tCre [mmol/kg] | 2 | 6.1 (0.41) | 6.2 (0.37) | $F$ = 2.5 | 0.118 | $\eta^2$ = 0.03 |
| tCho [mmol/kg] | 4 | 2.1 (0.18) | 2.1 (0.16) | $F$ = 0.0 | 0.962 | $\eta^2$ = 0.00 |
| tmI [mmol/kg] | 3 | 8.3 (0.70) | 8.4 (0.92) | $F$ = 0.5 | 0.488 | $\eta^2$ = 0.00 |
| tNAA [mmol/kg] | 3 | 8.5 (0.88) | 8.4 (0.63) | $F$ = 0.1 | 0.716 | $\eta^2$ = 0.09 |

Values are presented as mean (SD). *F*-statistics refer to linear models or Welch's tests for GABA and NAA due to inhomogeneity of variance. $\eta^2$ values refer to *partial* $\eta^2$'s.
[a]FDR-corrected for $n$ = 2 tests.
*n*-IC†: statistical inference changed with removal of influential cases.

**Supplementary Table 5 Associations of Glx/GABA and parallel CPM effects with clinical characteristics.**

| | Clinical characteristic | Test statistic | *P* | Missing values (*n*) |
|---|---|---|---|---|
| **Glx/GABA** | Average clinical pain intensity over past 4 weeks [NRS] | *rho* = -0.17 | 0.894[a] | 0 |
| | Pain duration [months] | *rho* = 0.05 | 0.965[a] | 1 |
| | Spatial pain extent [%] | *rho* = 0.12 | 0.894[a] | 0 |
| | Within-MRS-session clinical pain [NRS] | *rho* = 0.01 | 0.965[a] | 0 |
| **Parallel CPM effects** | | | | |
| CPM △PPT parallel LB [%] | Average clinical pain intensity over past 4 weeks [NRS] | *rho* = 0.29 | 0.185[b] | 1 |
| | Pain duration [months] | *rho* = -0.32 | 0.185[b] | 2 |
| | Spatial pain extent [%] | *rho* = 0.17 | 0.327[b] | 1 |
| | Within-CPM-session clinical pain [NRS] | *rho* = 0.27 | 0.190[b] | 1 |
| CPM △PPT parallel Hand [%] | Average clinical pain intensity over past 4 weeks [NRS] | *rho* = 0.54 | **0.003**[b] | 2 |
| | Pain duration [months] | *rho* = 0.10 | 0.545[b] | 3 |
| | Spatial pain extent [%] | *rho* = 0.18 | 0.327[b] | 2 |
| | Within-CPM-session clinical pain [NRS] | *rho* = 0.22 | 0.270[b] | 2 |

*Rho* statistics refer to Spearman correlations. Missing values were omitted from the analyses. For pain duration, 1 participant did not indicate the month of pain onset. For CPM, values are missing due to time constraints during the water bath or examiner error.
[a]FDR-corrected for *n* = 4 tests.
[b]FDR-corrected for *n* = 8 tests.

**Supplementary Table 6 Number of missing values for MRS outcomes, experimental pressure pain sensitivity and CPM effects.**

| | Missing values ($n$) | |
|---|---|---|
| | **CLBP patients ($n$ = 41)** | **Controls ($n$ = 29)** |
| SNR | | |
| FWHM $H_2O$ | | |
| FWHM NAA | | |
| Glx/GABA | | |
| Glx | | |
| GABA | | |
| tCre | 0 | 0 |
| tCho | | |
| tmI | | |
| tNAA | | |
| GM | | |
| WM | | |
| CSF | | |
| **Experimental pressure pain sensitivity** | | |
| PPT LB | 0 | 0 |
| PPT Hand | 0 | 0 |
| **CPM effects** | | |
| | **CPM-SHAM Controls ($n$ = 29)** | **CPM Controls ($n$ = 29)** |
| PPT before Hand | 0 | 1 |
| PPT during Hand | 0 | 1 |
| PPT after Hand | 0 | 0 |
| ∆PPT parallel Hand | 0 | 2 |
| ∆PPT sequential Hand | 0 | 1 |
| | **CLBP patients ($n$ = 41)** | **Controls ($n$ = 29)** |
| Cold-water bath LB pain intensity | 0 | 0 |
| Cold-water bath Hand pain intensity | 0 | 0 |
| PPT before LB | 0 | 0 |
| PPT during LB | 1 | 1 |
| CPM ∆PPT parallel LB | 1 | 1 |
| PPT before Hand | 0 | 1 |
| PPT during Hand | 2 | 1 |
| CPM ∆PPT parallel Hand | 2 | 2 |
| CPM-inhibitors LB | | |
| CPM-facilitators LB | 1 | 1 |
| CPM-non-responders LB | | |
| CPM-inhibitors Hand | | |
| CPM-facilitators Hand | 2 | 2 |
| CPM-non-responders Hand | | |

For Wilcoxon rank-sum tests, t-tests or linear models, missing values were omitted from the analyses. For CPM within-subject analyses, linear mixed models accounted for missing values using maximum likelihood estimation. CPM values are missing due to time constraints during the water bath or examiner error.
[a]Linear mixed model analysis.
